# Tumour-intrinsic features shape T-cell differentiation through myeloma disease evolution

**DOI:** 10.1101/2024.06.22.24309250

**Authors:** Kane A. Foster, Elise Rees, Louise Ainley, Eileen M. Boyle, Lydia Lee, Gwennan Ward, Daria Galas-Filipowicz, Anna Mikolajczak, Emma J. Lyon, Dylan Jankovic, Jasmine Rahman, Mahima Turakhia, Imran Uddin, Gordon Beattie, Yvette Hoade, Catherine Zhu, James L. Reading, Ieuan Walker, Michael Chapman, Karthik Ramasamy, Javier Herrero, Benny Chain, Sergio A. Quezada, Kwee L. Yong

## Abstract

The haematological malignancy multiple myeloma is associated with skewed T-cell activation and function. T-cell alterations are detectable in asymptomatic myeloma precursor conditions and have the potential to identify precursor patients at imminent risk of progression. However, what myeloma-associated T-cells alterations represent mechanistically, how they relate to tumour burden and gene expression, and what influences high inter-patient variability in immune composition remains unknown. Here, we assembled the largest ever dataset of published and newly-generated single-cell RNA and TCR sequencing of the marrow and blood from patients with myeloma, precursor conditions, and age-matched non-cancer controls. We show myeloma is not associated with T-cell exhaustion and instead defined by a pattern of T-cell differentiation resembling antigen-driven terminal memory differentiation. Myeloma-associated T-cell differentiation was dependent on tumour-intrinsic features including tumour burden and tumour expression of antigen-presentation genes. Expanded TCR clones accumulating in myeloma were not enriched for viral specificity and were detected in effector states in highly infiltrated marrows. Together, these results suggest anti-tumour immunity drives a novel form of cancer-associated T-cell memory differentiation in myeloma.

## Introduction

T-cells are polyfunctional immune cells and fundamental players in anti-tumour immunity^1^. In solid cancers, evidence suggests early in carcinogenesis tumour growth can be curtailed by tumour-reactive T-cells^1,2^. However, persistent activation drives these cells away from functional memory states towards a hypo-responsive state of terminal differentiation termed exhaustion^3,4^, characterised by the expression of immune checkpoint molecules like programmed cell death protein 1 (PD1)^5^, contributing to cancer progression in solid cancers. This complex interaction is believed to shape tumours from the early precursor stages to relapsed and refractory disease^1^. Understanding these insights have refined the treatment of solid tumours through the development of immunotherapies targeting exhausted T-cells^6^.

Multiple myeloma (MM) is a haematological malignancy of bone marrow (BM) plasma cells that is largely incurable^7,8^. Two precursor conditions of increasing severity precede MM: Monoclonal Gammopathy of Undetermined Significance (MGUS) and then Smouldering Multiple Myeloma (SMM). They differ in their risk of progression to overt myeloma with MGUS and SMM being associated with a 5-year progression rate of approximately 7% and 50% respectively^9^. While not every MGUS or SMM patient will progress, virtually every MM patient has transitioned through these stages^10^. Thus, there is a pressing clinical need to identify asymptomatic patients with precursor conditions at imminent risk of progression. Current risk factors rely largely on tumour bulk^11^, namely the levels of plasma cell infiltration in the bone marrow and serum concentrations of paraprotein (malignant cell-derived clonal immunoglobulin) and beta-2 microglobulin (B2m)^9^. However, the role of the BM tumour microenvironment, particularly T-cells, in progression remains poorly understood. Understanding how myeloma drives alterations in T cell state and function is complicated by the influence of patient advanced age and marrow homeostatic T-cell differentiation^7,12,13^. This, together with high inter-individual immune heterogeneity, confounds identifying associations between T-cells and tumour biology or progression.

To solve this, we combined over a million single cells from 295 samples from 237 donors using newly-generated single-cell RNA sequencing (scRNA-seq) and T-cell receptor (TCR) sequencing (scTCR-seq) data and 11 published studies^14,15,24,16–23^, allowing us to interrogate T-cell dynamics while controlling for natural and tumour-associated sources of inter-individual variation. We show for the first time that the T-cell landscape associated with myeloma possess features of antigen-driven terminal memory differentiation and highlight the features of tumour biology driving this. These results suggest that anti-tumour immunity underpins a novel form of tumour-associated T-cell differentiation in myeloma.

## Results

### Effective integration of scRNA-seq datasets allows a detailed classification of immune cell populations

To study immune differentiation through myeloma disease evolution we generated a large scRNA-seq map of BM and peripheral blood (PB) cells from untreated MGUS (n = 20, 9%), SMM (n = 58, 25%) and MM (n = 54, 23%) patients alongside non-cancer controls (n = 102, 44%; Fig. 1a, Extended Data Fig. 1a, Supplemental Table 1). Patients were older than controls (controls median 55 range 21–87, MGUS median 62 range: 41–81, SMM median 62 range: 29–81, MM median 62 range: 38–77; Fig. 1b). As expected, plasma cell infiltration of the bone marrow and serum paraprotein levels rose with from SMM to MM (infiltration *P* = 0.001, paraprotein *P* < 0.05, Wilcoxon test; Fig. 1c).

**Fig. 1.**
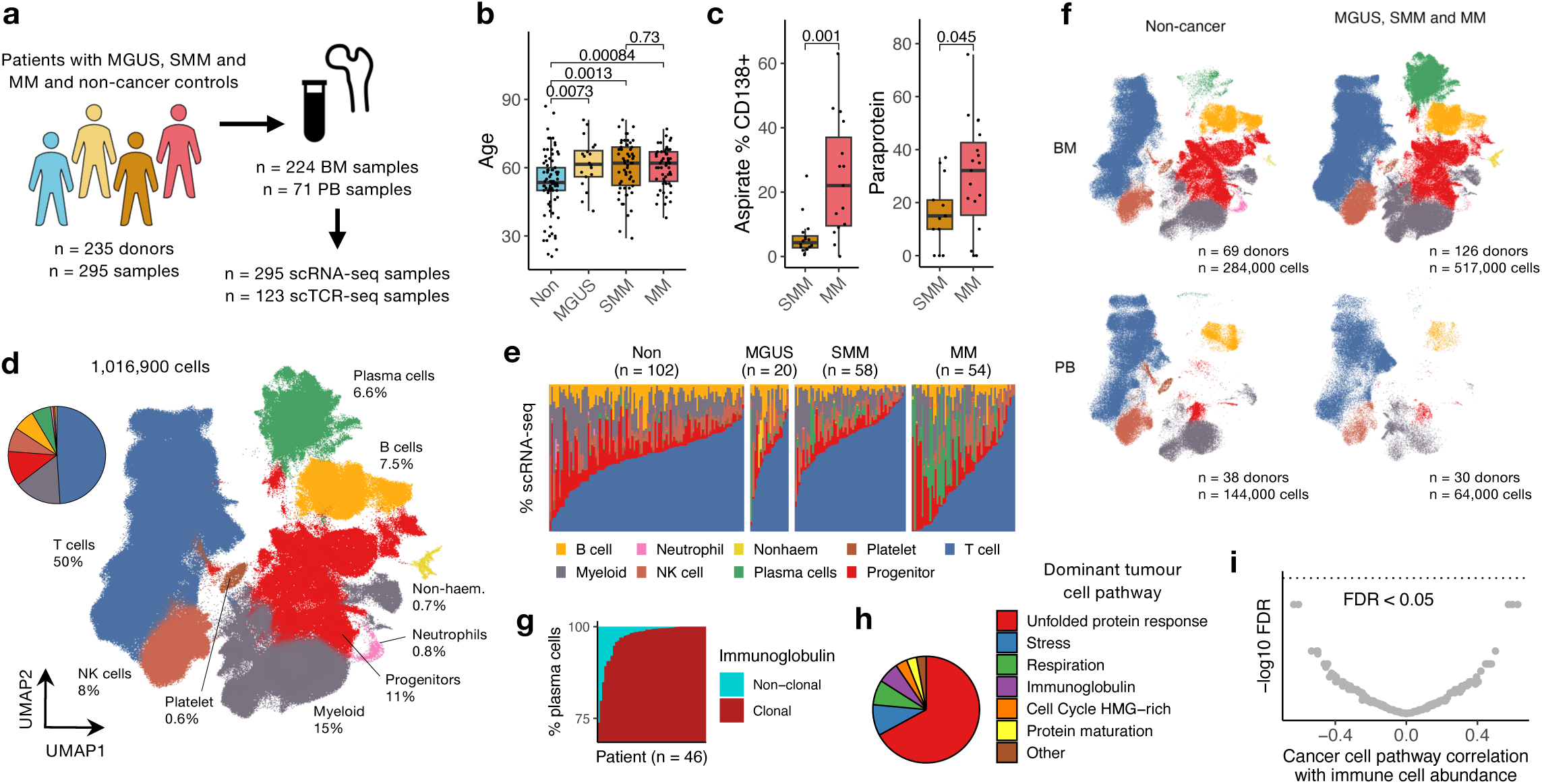
| Single cell phenotypes are harmonized in a large integrated scRNAseq dataset of patients with myeloma and precursor conditions and non-cancer controls. **a**, Schematic depicting the cohort, tissue types (BM, bone marrow; PB, peripheral blood), and singe-cell data types included in the study. **b**, Box plot showing the distribution of age (years) in non-cancer controls (n = 86), MGUS (n = 20), SMM (n = 58) and MM (n = 53) patients. **c**, Box plots showing the distribution of aspirate % CD138+ (left) and paraprotein values (right) in SMM (n = 13) and MM (n = 19) patients. **d**, Visualisation of all cells in the dataset by uniform manifold approximation and projection (UMAP). The colour each point (cell) represents the indicated cell type cluster. The percentage of all cells occupied by each cluster is inset. **e**, Bar chart showing the proportion of each cluster for each individual donor (columns) for each cohort. **f**, UMAP as in (**d**) separated by cohort (columns) and tissue type (rows). The number of donors and cells for each separation is inset. **g**. Bar chart showing the proportion of plasma cells with clonal or non-clonal immunoglobulin usage. Plasma cells with clonal immunoglobulin usage were classified as tumour cells. **h**. Pie chart quantify the proportion of each pan-cancer transcriptional pathway which was the most highly expressed (dominant) in tumour cells across all patients. **i**, Dot plot showing FDR-adjusted *P* values and correlation coefficients between cell type cluster abundance (as a proportion of non-plasma cells) and pan-cancer transcriptional pathway expression in tumour cells (n = 45 patients). Box plots represent the first and third quartiles around the median with whiskers extending 1.5 times the interquartile range. *P* values shown on box plots were calculated by two-sided Wilcoxon test.

Following quality control and correcting for batch effects (see Methods), Extended Data Fig. 1b), cells were clustered to 9 major cell types and phenotyped using RNA expression, protein expression via cellular indexing of transcriptomes and epitopes (CITE-seq), and *de novo* label prediction tools (Fig.1d, Extended Data Fig. 1c,d). T-cells (defined by co-expression of *CD3D, CD3E, CD3G*, *CD8A* and *CD4* RNA and CD3 protein) comprised roughly half (50.2%) the cells in the dataset (Fig. 1e), with another quarter occupied by similar proportions of myeloid cells (*FCN1*+*FCER1G*+CD14+; 15%) and haematogenic progenitors (*CD34*+*MPO*+*TYMS+*; 11%). The remainder of the dataset was comprised of equivalent numbers of NK cells (*KLRD1*+*FCG3RA*+CD56+; 8%), B cells (*CD79A*+CD19+; 7.5%), and plasma cells (*MZB1*+*SDC1+*; 6.6%), alongside small (<1% total counts) clusters of neutrophils (*NEAT1*+*NAMPT*+), non-haemopoietic cells (*CXCL12*+*COL3A+*), and platelets (*PPBP*+*PF4+*).

Despite the heterologous sorting strategies employed by different studies (Extended Data Fig. 1b), we identified major determinants of cellular composition in our dataset (Fig. 1f). Plasma cells and progenitors were enriched in the BM relative to PB (*P* < 0.001 and *P* < 0.001, respectively, Wilcoxon test), suggesting a relative lack of haemodilution in BM aspirates. As expected, plasma cells were most highly enriched in the BM of patients relative to controls (*P* < 0.001, Wilcoxon test). However, the global distribution of cell types was otherwise similar in diseased and controls marrows, suggesting progression to myeloma may be associated with more granular alterations to immune composition.

Overall, we analysed 1,009,317 cells from 234 individuals with RNA and clinical data, 109 with TCR and 1 with CITE-seq data, including 224 BM and 71 PB samples. This resource represents a large cross-sectional analysis of controls and myeloma disease stages and reliably discriminate key immune phenotypes of non-cancer controls and myeloma patients through disease evolution.

### Recurrent transcriptional pathways in malignant plasma cells underpin progression and outcome

Next, we sought to characterise more granular features associated with disease evolution that may reveal tumour-immune cross-talk by conducting an analysis of tumour cells. Malignant clones, identified by clonal immunoglobulin usage (see Methods; Extended Data Fig. 2a-c), composed the majority of plasma cells in all patients but were most abundant in MM (Fig. 1g and Extended Data Fig. 2f). To overcome inter-patient tumour transcriptional heterogeneity we scored 67,656 plasma cells from 46 patients with a recently published set of pan-cancer transcriptional pathways^25^ (Extended Data Fig.2e, Supplemental Table 2). The pathway most highly expressed by tumour cells corresponded to the unfolded protein response (67% tumour cells, Fig. 1h), with the remaining cells defined by pathways reflecting cellular stress, respiration, or other biological processes.

To explore the significance of these pathways in disease evolution, we compared their expression in SMM (n = 17) and MM (n = 29; Extended Data Fig. 2f-g), analysed associations with local marrow infiltration (Extended Data Fig. 2h), and overall survival in MM patients from the CoMMpass study^26^ (Extended Data Fig. 2i). We identified 18 pathways significantly associated with progression or marrow infiltration, of which 6 were associated with outcome. Broadly, pathways reflecting proteostasis such as the unfolded protein response were characteristic of SMM (*P* < 0.001, Wilcoxon test), whereas more functionally diverse set of pathways were enriched at progression (Extended Data Fig. 2f-g). Proliferation pathways were enriched at progression (*P* = 0.07, Wilcoxon test), in highly infiltrated marrows (*R* = 0.42, *P* = 0.03, Pearson correlation), and associated with significantly shorter overall survival (*P* < 0.001, log-rank test; Extended Data Fig. 2i). Conversely, a SMM-enriched pathway characterised by immunoglobulin genes was more frequently highly-expressed in less-infiltrated marrows and association with more favourable outcome (*P* < 0.001, log-rank test). This suggests plasma cells retaining normal functions, such as proteostasis^27^, were more common in low risk-and low burden disease, whereas more proliferative states are associated with worse outcome.

Cellular stress was among the pathways more frequently expressed in MM (P = 0.007, Wilcoxon test; Extended Data Fig. 2e). We extricated this pathway from *in vitro* stressors such as those introduced by sample processing (Extended Data Fig. 2j). Closer inspection of tumour cells highly expressing the stress pathway revealed an enrichment of genes also associated with cell death (*P* = 0.02, GSEA of programmed cell death pathway; Extended Data Fig. 2k). Death pathways were enriched at progression and positively correlated with tumour burden (Extended Data Fig. 2k, Supplemental Table 2), suggesting this stress pathway may reflect death-associated processes. Despite an enrichment at progression, high expression of the stress pathway was associated with superior outcome (*P* = 0.014, log-rank test; Extended Data Fig. 2i), suggesting a more complex relationship for this pathway through disease evolution possibly related to immune correlates.

Finally, we compared tumour pathways with immune composition. Tumour cell pathway expression was not associated with the abundance (as a fraction of non-plasma cells) of cell types in the BM (all individual cell type–tumour pathway correlations adjusted *P* > 0.05, Pearson correlation; Fig. 1i).

Together, this data show how the transcriptional activity of tumour cells is influenced related to progression, infiltration, and outcome. However, connecting these pathways to tumour– immune interactions may require a more granular overview of immune cells.

### In-depth T-cell phenotyping reveals myeloma is not enriched in exhausted cells and T-cell composition is similar in health and disease

To more deeply probe immune perturbations and tumour-immune cross-talk in disease evolution, T-cells were isolated, re-integrated and re-clustered to 19 discrete phenotypes and transcriptional states based on expression of canonical RNA and protein markers (Fig. 2a-b, Extended Data Fig.3a-c, Supplemental Tables 2-3)^4,28–30^. CD4+ cells were predominantly naïve (Tn, 49% of CD4+T-cells) and central memory (Tcm, 21%) cells, and the remainder in regulatory (Treg, 8.4%), helper (Th17 7.2%; T effector memory, Tem 6.1%) or cytotoxic (cytotoxic T lymphocyte, CTL, 8.7%) states. CD8+ cells were classified into a more linear trajectory spanning earlier Tn (20% of CD8+T-cells), Tcm (4.6%) and *IL7R*-expressing effector memory (Tem.IL7R, 12%) through *PDCD1*+ activated Tem (TemActive, 22%) to more later differentiated *GZMB*+ terminal Tem (TemTerm, 18%) and Tem re-expressing CD45RA (TEMRA, 15.1%) subsets, alongside tissue resident (Trm, 5.1%) and exhausted (Tex, 0.7%) clusters. Invariant subsets, comprising γδ T-cells (gdT) and mucosal-associated invariant T-cells (MAIT), proliferating and interferon-induced clusters were also identified. This included an interferon-induced cluster resembling effector T-cells characterised by expression of IFN-induced protein with tetratricopeptide repeats 2 (*IFIT2*) alongside effector molecules *TNF* and *IFNG* (Teff.IFIT2). This functional annotation was consistent with patterns of TCR expansion (Fig. 2c, Extended Data Fig. 3d-e), with the most clonally expanded clusters expressing the highest level of late differentiation markers. Our phenotypes showed high concordance with published and predicted cluster labels (Extended Data Fig. 3f), arguing for a faithful representation of T-cell phenotypes in our integrated dataset.

**Fig. 2.**
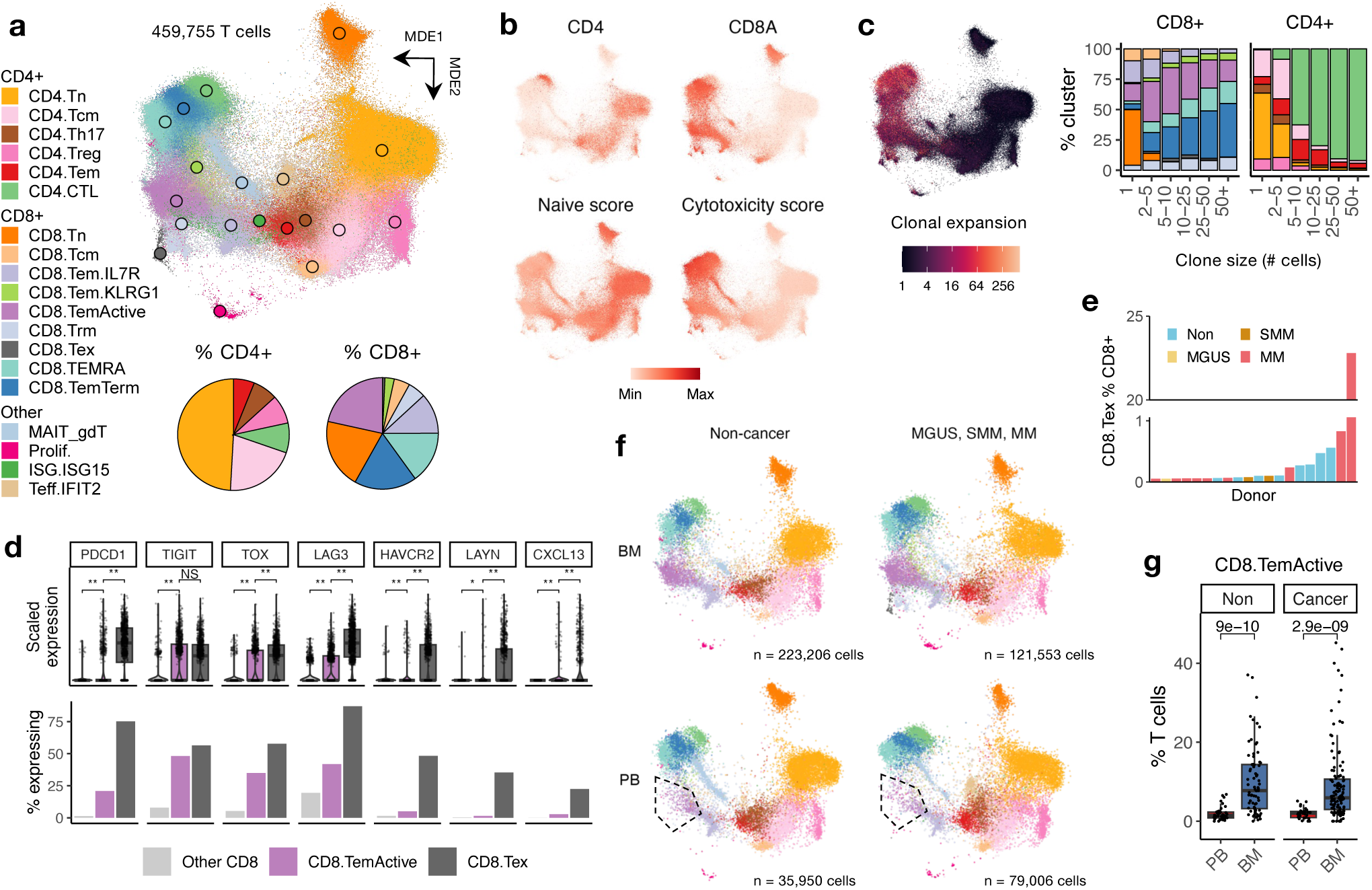
| The T cell landscape of myeloma is not enriched in exhausted cells and reflects intrinsic marrow biology. **a**, Visualisation of all T cells in the dataset by minimum-distortion embedding (MDE). The colour each point (cell) represents the T cell cluster indicated by the legend. Pie charts depict the percentage of CD4+ (left) and CD8+ (right) T cell clusters in the dataset. **b**, MDE plots showing the expression of *CD4*, *CD8A*, and naïve and cytotoxic genes signatures. **c**, Left, MDE plot showing the clonal expansion of T cell receptor (TCR) clones (calculated as the number of times a unique clone was seen). Right, Bar chart showing the proportion of each CD8+ (left) and CD4+ (right) cluster among TCR clones of the indicated size. Bar colour represents T cell cluster indicated in (**a**). **d**, Box plots showing the expression (top) and bar plots showing the percentage of cells expressing (bottom, fraction of cells with non-zero expression) indicated T cell exhaustion associated genes in CD8.Tex, CD8.TemActive and other CD8+ T cell clusters. For the three groups a random sample of 1,000 cells from Zheng et al. are shown. *P* values calculated by Wilcoxon test, ** = *P* < 0.001, * = *P* < 0.01, NS = *P* > 0.05. **e**, Bar plot showing the abundance of CD8.Tex as a percentage of total CD8+ T cells for the twenty donors with the highest abundance of CD8.Tex. **f**, MDE as in (**a**) separated by cohort (columns) and tissue (rows: bone marrow, BM; peripheral blood, PB). For the four groups a random sample of 20,000 cells is shown. The total number of cells for each separation is inset. The CD8.TemActive cluster is circled. **g**, Box plot showing the abundance of CD8.TemActive as a percentage of T cells in the PB and BM of noncancer controls (Non, PB n = 39, BM n = 73; left) and SMM and MM patients (Cancer, PB n = 30, BM n = 123; right). Box plots represent the first and third quartiles around the median with whiskers extending 1.5 times the interquartile range. *P* values shown on box plots were calculated by two-sided Wilcoxon test.

We validated our proposed BM T-cell landscape by employing CyTOF on an independent cohort of 9 SMM and 11 MM donors, assaying 940,000 cells with 46 markers (Supplemental Table 4). *De novo* clustering and a comparison of T-cell clusters across technologies (see Methods) revealed a range of phenotypes closely matched and enhanced those seen in our scRNA-seq dataset (Extended Data Fig. 4). For example, expression of the CD57 glycoepitope on GZMB+ CD28– CD8+Tem (CD8.Tem-Term) suggested a phenotype of terminal effector memory cells^31,32^.

We identified exhausted CD8.Tex by the expression of high levels of *PDCD1* and *TIGIT*, alongside other RNA markers of exhaustion like *CXCL13* and *LAYN*^29^. Importantly, we distinguish CD8.Tex from *GZMK*-expressing CD8.TemActive. CD8.TemActive expressed higher *PDCD1* and *TOX* than non-exhausted bone marrow T-cells (*P* < 0.001 and *P* < 0.001, Wilcoxon test) but less than CD8.Tex (*P* < 0.001 and *P* < 0.001, Wilcoxon test) and lacked other markers of exhaustion like *LAYN* (*P* < 0.001, Wilcoxon test; Fig. 2d, Extended Data Fig. 3b) and expressed early differentiation markers like *CD28*. In our CyTOF dataset, expression of PD1 and the exhausted-associated transcription factor TOX^33^ were similarly restricted to early CD8+Tem (CD45RO+KLRG1+CD28+; Extended Data Fig. 4c). Interestingly, the CD8.Tex cluster was almost entirely composed of cells from a single myeloma patient (1181 of 1222 cells, 97%; Fig. 2e) who contributed the majority of exhaustion marker-expressing cells (Extended Data Fig. 2g), suggesting CD8.Tex were a donor-specific phenomenon. Similar observations were made for the CD8.Trm cluster, being mostly composed of 2 samples from the same study as the donors-specific CD8.Tex (27.4% and 26.1% of cells; Extended Data Fig. 5a). These data lead us to suggest that exhausted T-cells are rarely seen in myeloma and the more frequent PD1-expressing activated CD8+Tem are distinct from exhausted cells.

Similar T-cell phenotypes were observed in the BM and PB (Fig. 2f), but the proportion of T-cell clusters differed between tissues (Extended Data Fig. 3h). Notably, CD8.TemActive were enriched in the BM of both patients and controls (*P* < 0.001 and *P* < 0.001, Fig. 2g), suggesting intrinsic marrow biology regulates the abundance of this subsets in health and myeloma.

### T-cell differentiation skewing occurs in disease independent of age and presents similarly in pre-malignant SMM and overt MM

We next asked how the relative abundance of T-cell subsets in the BM was altered across disease stages. BM T-cell composition was strikingly similar in patients and controls (Fig. 3a, Extended Data Fig. 5a), with the exception of donor-specific CD8.Tex clusters. We next compared the T-cell composition of controls with each myeloma disease stage in turn, statistically controlling for age. The most prominent difference in BM T-cell composition between health and disease was the loss of naïve, CD4.Th17 and MAIT cells and an enrichment of *GZMB*-expressing memory T-cell clusters (FDR-adjusted *P* < 0.1 for all, linear models; Fig. 3b, Extended Data Fig. 5b). When removing the one patient who contributed the majority of CD8.Tex cells (97% cells; Fig. 2e), this cluster was not enriched in MM relative to controls (*P* = 0.54, linear models). As non-cancer controls included hip replacement and deceased donors, we repeated our analysis with only healthy donors and obtained the same results (Extended Data Fig. 5c). Low-risk MGUS possessed a T-cell composition with the fewest differences to control marrows. Conversely, T-cell composition was similar between the higher-risk but pre-cancerous SMM and symptomatic MM (Extended Data Fig. 5d). While the normalised abundance of CD8.Tex was lower in SMM than MM independent of age (*P* < 0.001, linear model), in terms of unnormalized counts this only represented 8 MM patients with a median of 1 CD8.Tex cells each suggesting this did not represent a meaningful enrichment. For patients with available risk data, we saw no significant differences in T-cell composition between international staging system (ISS) and SMM Mayo risk groups^9,34^ (Extended Data Fig. 5e). In our smaller CyTOF cohort we noted a trend for the enrichment of CD57+ CD8.Tem-Term in MM relative to SMM (Extended Data Fig. 5f). These results suggest that smouldering and overt myeloma are associated with similar T-cell alterations independent of age.

**Fig. 3.**
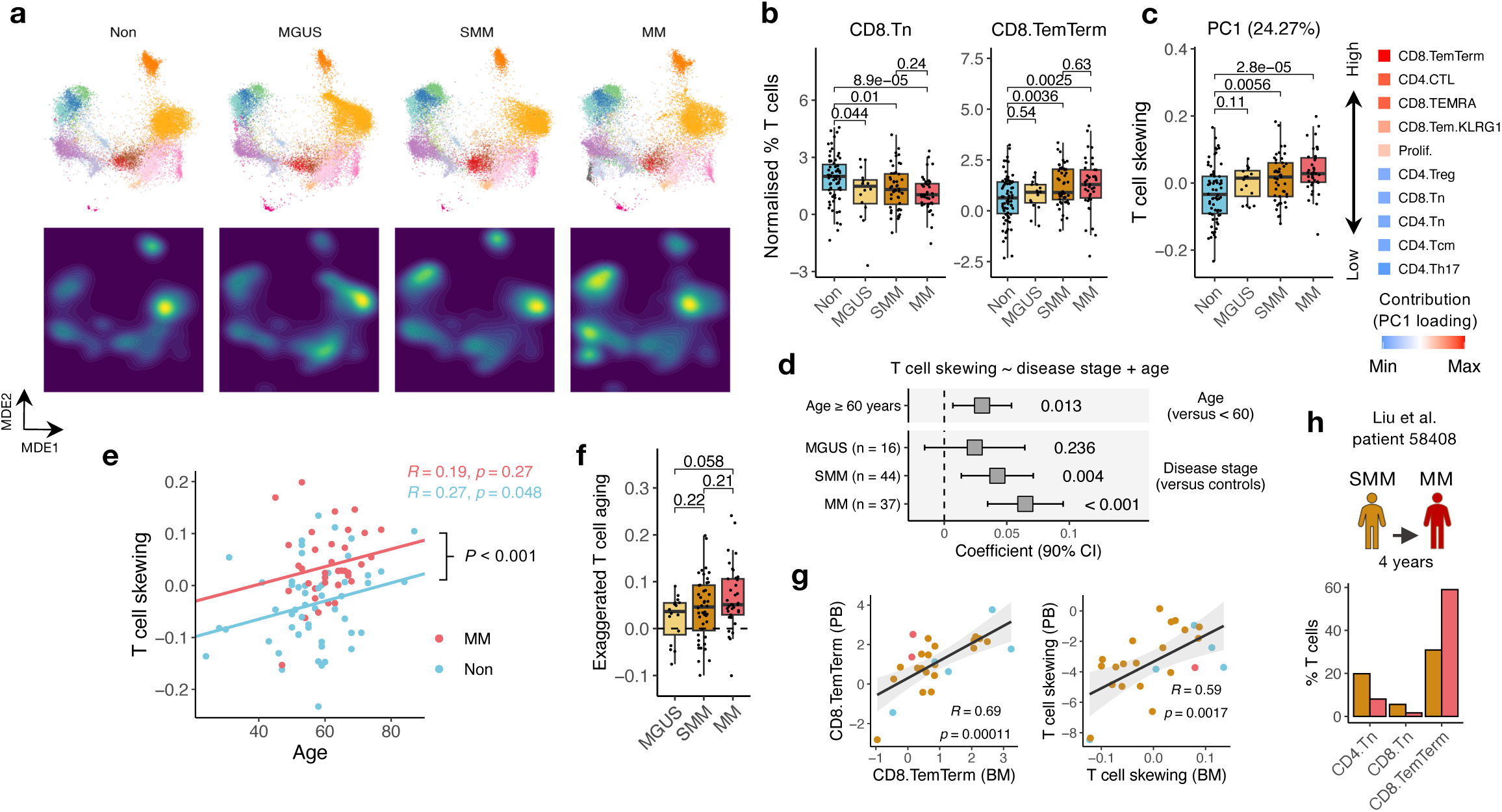
| Step-wise alterations to bone marrow T cell composition occur through myeloma disease evolution. **a**, Visualisation of BM T cells clusters (top) and cell density (bottom) by minimum-distortion embedding (MDE) in non-cancer controls (Non), MGUS, SMM and MM patients. For the four groups a random sample of 20,000 cells is shown. **b**, Box plots showing the normalised abundance (see Methods) of CD8.Tn (left) and CD8.TemTerm (right) in the BM as a percentage of T cells in non-cancer controls (n = 71), MGUS (n = 16), SMM (n = 48) and MM (n = 41) patients. **c**, Left, Box plot showing the value of the T cell skewing (the first principal component (PC1), 24.27% variance, calculated on BM samples only) in Non (n = 68), MGUS (n = 16), SMM (n = 45) and MM (n = 39) patients. Right, representation of the clusters with the highest and lowest contribution (loading) to PC1. For example, a high PC1 value corresponds to a high number of CD8.TemTerm. **d**, Forest plot showing the relationship between T cell skewing and disease stage (MGUS (n = 16), SMM (n = 44) and MM (n = 37) relative to controls (n = 53)) and age (binarized to ≥ median age (60 years)). Coefficients with 90% confident interval (CI) and *P* values from linear model (see Methods) are inset. **e**, Dot plot showing the correlation between T cell skewing and age in non-cancer controls and MM patients. *P* value from linear model is inset. **f**, Box plot showing exaggerated T cell aging (the residuals between a patient’s PC1 values and a model of PC1 and age in controls only, see Methods) in MGUS, SMM and MM patients. Residuals of zero (T cell skewing expected for patient’s age) is indicated with dashed line. **g**, Dot plots showing the correlation between the abundance of CD8.TemTerm (left) and T cell skewing (right) in the PB and BM for patients with samples from both tissues. **h**, Upper, Schematic depicting longitudinal sampling of patient 58408 from Liu et al. Lower, Bar plot showing the abundance of indicated T cell cluster in SMM and MM for patient 58408. Box plots represent the first and third quartiles around the median with whiskers extending 1.5 times the interquartile range. *P* values shown on box plots were calculated by two-sided Wilcoxon test. R and *P* values for correlations were calculated by Pearson correlation. Correlation shaded regions represent the 95% confidence interval of linear regression slopes.

**Fig. 4.**
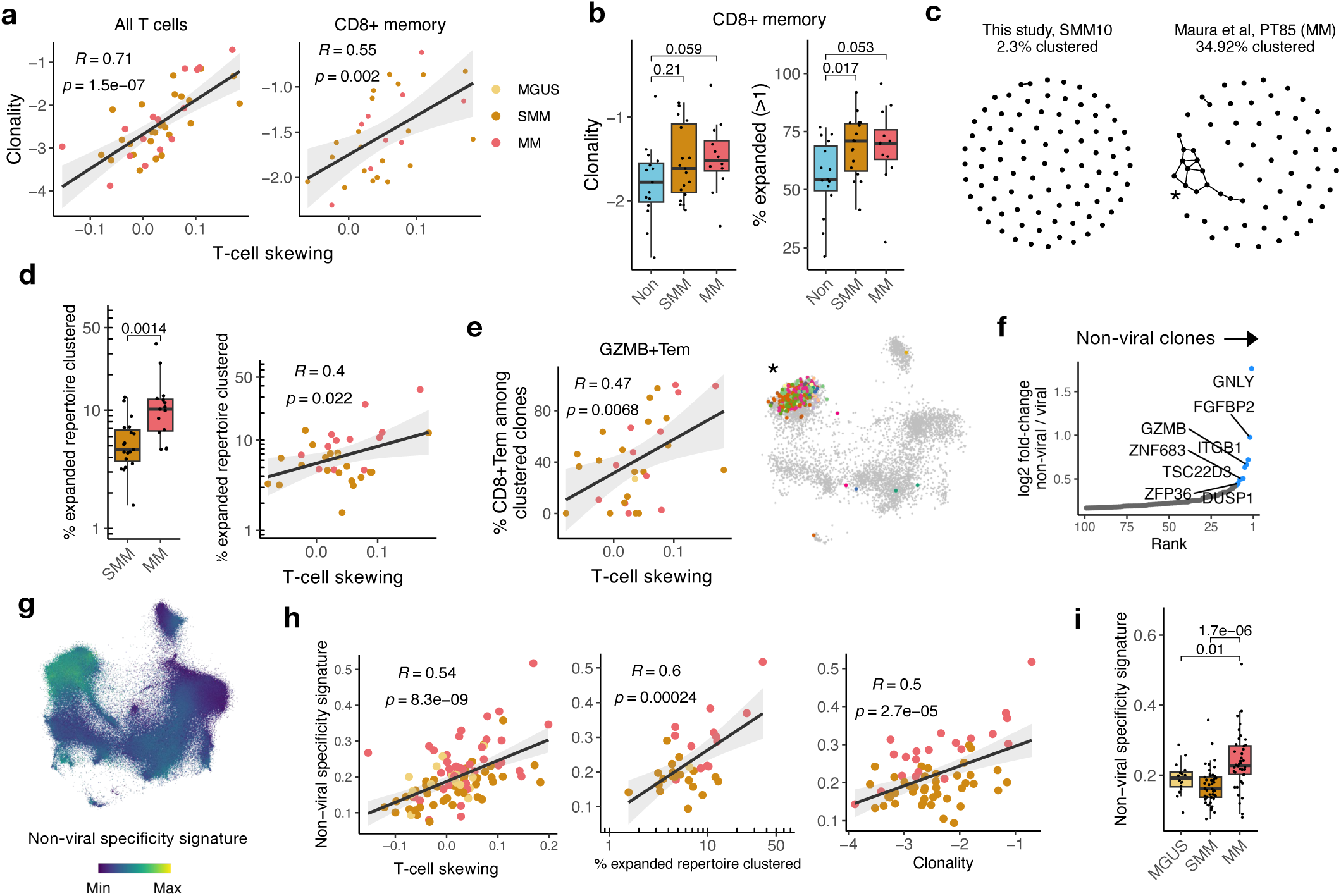
| Features of antigen-experienced T cell receptor repertoires underpin myeloma-associated T cell differentiation. **a**, Dot plots showing the correlation between T cell skewing (PC1 values) and TCR clonality (log_10_ 1/Simpson’s diversity) among all T cell clones (left) and CD8+ memory clones (CD8+ clones, CD8+ clusters excluding CD8.Tn and CD8.Tcm) in MGUS, SMM and MM patients (all T cell clones n = 42, CD8+ memory clones n = 29). **b**, Box plots showing the clonality (left) and abundance of expanded clones (right) of CD8+ memory clones in non-cancer controls (n = 15), SMM (n = 19) and MM (n = 12). **c**, Network plots showing the extent of clustering among expanded (>1) TCRs from two representative patients with low (left) and high (right) percentages of total repertoire clustering. Each node represents a TCR clone and each edge that two TCR clones had a co-clustered alpha or beta sequence. An asterisk indicates a cluster analysed in **e**. **d**, Left, Box plot showing the percentage of clustered expanded TCRs in SMM (n = 19) and MM (n = 15). Right, dot plot showing the correlation between PC1 values and the the percentage of clustered expanded TCRs in MGUS, SMM and MM patients (n = 35). **e**, Left, Dot plot showing the correlation of T cell skewing and the percentage of CD8+Tem (CD8.TemTerm, CD8,TEMRA) among cells from clustered TCR clones. **f**, Results from a differential expression analysis of T cells possessing TCR clones annotated as viral-reactive (see Methods) versus all other clones. Expression testing was performed in 19 patients with a median of 20 viral-reactive and 966 unannotated clones per-patient. The 10 most highly non-viral enriched genes were used to define a non-viral specificity signature and are labelled. **g**, Visualisation of 1,016,900 T cells by minimum-distortion embedding (MDE). The colour each point (cell) represents the non-viral specificity signature score expressed by that T cell. **h**, Dot plots showing the correlation between the mean non-viral specificity signature score per-patient and T cell skewing, the percentage of clustered TCRs among expanded TCRs, and TCR clonality in MGUS, SMM and MM patients. **i**, Box plot showing the mean non-viral specificity signature score in MGUS (n = 16), SMM (n = 45) and MM (n = 40) patients. Box plots represent the first and third quartiles around the median with whiskers extending 1.5 times the interquartile range. *P* values shown on box plots were calculated by two-sided Wilcoxon test. R and *P* values for correlations were calculated by Pearson correlation. Correlation shaded regions represent the 95% confidence interval of linear regression slopes.

To interrogate changes to BM T-cells in a less supervised manner we ran principal component analysis (PCA) on patient’s T-cell composition. The first principal component explaining the highest fraction of variance in T-cell composition (PC1, 24.27%; Extended Data Fig. 5g) described a compositional shift across clusters from more naïve and early subsets to terminal memory clusters (Fig. 3c). As this composition alteration represented a shift from phenotypes at either end of the T-cell differentiation spectrum^4^, we termed PC1 “T-cell skewing”. T-cell skewing was highest (indicating an enrichment of terminal memory clusters) in SMM and MM relative to controls independent of age (*P* < 0.004 and *P* < 0.001, respectively, linear model; Fig. 3c-d), demonstrating this metric captured the major alterations to T-cells in myeloma. We noted T-cell skewing was associated with age independent of patient group (*P* = 0.013, linear model, R = 0.28, Pearson correlation; Fig. 3d, Extended Data Fig. 5h) and correlated with age in controls (*P* < 0.05, R = 0.25; Fig. 3e). As T-cell skewing was independently associated with both myeloma and age (Fig. 3d), this component of myeloma-associated T-cell differentiation resembled T-cell alterations seen during aging. Therefore, patients possessed an exaggerated form of the T-cell compositional skewing seen with aging. The degree of exaggerated T-cell aging (see Methods) trended to rise with disease severity (MGUS versus MM, *P* = 0.06, Wilcoxon test; Fig. 3f) and the highest fraction of patients with exaggerated skewing was seen in myeloma (86%) versus MGUS (68%) and SMM (72%).

PC1 values and the abundance of terminal memory subsets in the BM strongly correlated in the PB of the same patients (Fig. 3g), further indicating a similarity of these changes to systemic T-cell alterations seen with aging^12^.

Finally, in a single patient sampled longitudinally at SMM and at progression to MM we observed the same compositional alternations seen cross-sectionally (Fig. 3h), suggesting these differentiated T-cell phenotypes accumulate longitudinal within patients through disease evolution.

### Features of antigen-specific responses underpin myeloma-associated T-cell differentiation

Next, we analysed features of the TCR repertoire. Repertoire clonality was associated with T-cell skewing in patients independent of age (*P* < 0.001, linear model, R = 0.71; Fig. 4a). We observed similar results when restricting analysis to CD8-expressing memory clones (*P* < 0.01, linear model, R = 0.55; Fig. 4a, Extended Data Fig. 6a), with both the clonality and the abundance of expanded clones of this subset was trending for enrichment in MM relative to controls (Fig. 4b). T-cell skewing and CD8+ memory diversity did not correlate in controls (*P* = 0.66, linear model; Extended Data Fig. 6b), suggesting enhanced clonal expansion may be a unique feature T-cell differentiation in myeloma.

The accumulation of TCRs possessing similar CDR3 sequences can indicate responses against shared antigens^35^. Using tcrdist3^36^ we grouped all expanded TCRs in the dataset (11,545 clones) into 279 clusters (composed of 1,014 clones, 8.8% of input; Extended Data Fig. 6c-d, Fig. 4c). This analysis revealed an increasingly large fraction of the TCR repertoire was occupied by clustered clones in MM (median 8.7% range 3-35) relative to SMM (median 3.1% range 0-11.3) and this fraction correlated with T-cell skewing (R = 0.4, *P* = 0.02, Pearson correlation; Fig. 4d). T-cell skewing was specifically associated with the clustering of *GZMB*-expressing CD8+ memory cells (R = 0.47, *P* = < 0.01; Fig. 4e), suggesting conserved antigen-specific responses drive T-cell differentiation in myeloma, specifically among *GZMB*-expressing subsets.

We next explored the T-cell antigen specificity. TCR specificity databases are mostly composed of viral antigens^37–39^, allowing us to ask if viral antigen specificities were involved in T-cell differentiation and clonality in myeloma (see Methods). In 19 patients we identified putative HLA-matched specificities for a median of 7 (range 2-88) paired clone per-patient against viral antigens (Extended Data Fig. 6e). Comparing gene expression between clones with and without putative viral-specificity annotations, we observed clones predicted to be non-viral specific expressed genes characteristic of terminal memory such as *GZMB*, perforin/*PRF1* and Hobit/*ZNF683* (Fig. 4f, Supplemental Table 5). We summarised the expression of these genes into a non-viral specificity gene signature (Extended Data Fig. 6f). Non-viral specificity mapped to *GZMB*+ terminal memory clusters (Fig. 4g, Extended Data Fig. 6g), correlated with T-cell skewing, clonality and repertoire clustering (Fig. 4h), and was enriched in MM relative to precursor conditions (Fig. 4i). Together, these data show myeloma-associated T-cell differentiation occurs alongside alterations to the TCR repertoire resembling antigen-direct T-cell immunity which may not reflect the activity of viral-specific TCR clones.

### Tumour-intrinsic features drive two clonally-related patterns of T-cell differentiation

We next asked if features of tumour biology may drive T-cell skewing. We examined serum paraprotein and B2m concentrations, plasma cell marrow infiltration, and tumour cell transcriptional state (Extended Data Fig. 2). As T-cell skewing overlapped in precursor patients and overt MM (Fig. 3c), we looked for associations among all patients independent of disease stage.

We did not find an association between T-cell skewing and tumour transcriptional state, marrow infiltration, or B2m levels (Fig. 5a, Extended Data Fig. 7a-b). However, we observed a positive correlation between high T-cell skewing and an enrichment of terminal memory cells with paraprotein levels (*R* = 0.45, *P* = 0.02, Pearson correlation; Fig. 5a-b, Extend Data Fig. 7c). This association was seen across different disease stage and centres (Extended Data Fig. 7d) and was recapitulated in our CyTOF cohort (Extended Data Fig. 7e). The same paraprotein correlation was also seen with TCR repertoire clonality (*R* = 0.53, *P* < 0.001, Pearson correlation; Fig. 5a, Extended Data. 7f). We found the degree of exaggerated T-cell aging was significantly higher in SMM and MM patients with high paraprotein levels independent of disease stage (*P =* 0.01, linear model; Extended Data Fig. 7g), suggesting paraprotein was the main driver of T-cell skewing in patients.

**Fig. 5.**
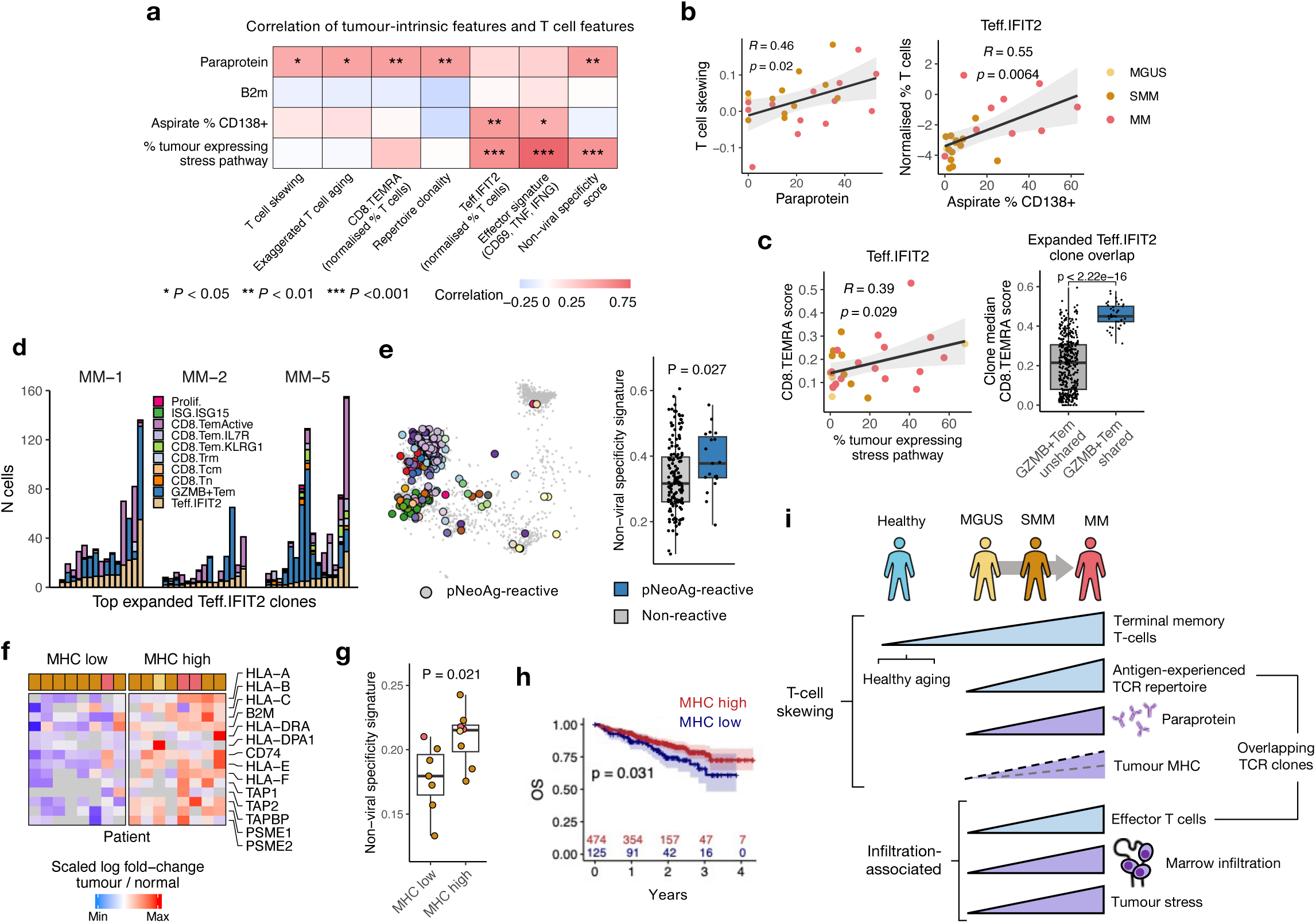
| Tumour-intrinsic drivers of T cell differentiation indicate anti-tumour immunity contributes to myeloma-associated T cell differentiation. **a**, Heatmap showing the correlation between tumour-intrinsic features (rows) and T cell features (columns) in all patients. Cell colour represents strength of association (correlation) and asterisk degree of significance (Pearson correlation). **b**, Dot plot showing the correlation between T cell skewing (PC1) and paraprotein values (left) and the normalised abundance (see Methods) of Teff.IFIT2 and the aspirate % CD138+ (right). **c**, Left, correlation between the average expression the twenty most significant CD8.TEMRA marker genes among Teff.IFIT2 cells and the the abundance of cancer cells highly expressing stress pathway genes. Right, Box plot showing the median expression of the CD8.TEMRA marker gene score per-clone among cells in the Teff.IFIT2 cluster. Each clone was grouped based on the T cell cluster composition of the remaining cells that composed that clone, specifically the abundance of *GZMB*-expressing Tem clusters (CD8.TEMRA, CD8.TemTerm and CD4.CTL), classifying those which were >50% composed of *GZMB*-expressing clusters as Shared and that those which were not as Unshared. **d**, Bar plot showing the T cell cluster composition of the largest CD8+ clones expanded in the Teff.IFIT2 cluster in three representative myeloma patients. Expanded Teff.IFIT2 clones are predominantly in GZMB+Tem (CD8.TEMRA, CD8.TemTerm and CD4.CTL) clusters. **e**, Left, Visualisation of T cells by minimum-distortion embedding (MDE). The coloured points (cell) represents cells derived from TCR clones predicted as reactive against cancer cell-expressing neoantigens (pNeoAg-reactive, see Methods). Point colour reflects individual clones. Right, Box plot showing the median expression of the non-viral specificity signature in expanded clones annotated as either pNeoAg-reactive or with no annotation. **f**, Heatmap showing the expression of genes significantly up– and down-regulated on cancer cells relative to non-cancer plasma cells across patients. Patients were classified as “MHC high” if their tumour cells had significantly upregulated the antigen processing and presentation pathway relative to nontumour cells and as “MHC low” if not. **g**, Box plot showing the expression of the non-viral specificity signature in patients with high and low expression of MHC genes (**f**). **h**, Kaplan–Meier curve showing the impact of high (red) and low (blue) scoring of the MHC pathway (**f**) on overall survival in 598 newly-diagnosed untreated multiple myeloma patients enrolled in the ComMMpass trial. *P* value calculated using log-rank test. **i**, schematic representing a summary of findings. Box plots represent the first and third quartiles around the median with whiskers extending 1.5 times the interquartile range. *P* values shown on box plots were calculated by two-sided Wilcoxon test. R and *P* values for correlations were calculated by Pearson correlation. Correlation shaded regions represent the 95% confidence interval of linear regression slopes.

We speculated tumour-intrinsic features were associated with individual T-cells clusters independent of overall T-cell skewing. Analysis of T-cell cluster abundance with marrow infiltration and transcriptional pathway scores in tumour cells revealed effector-like Teff.IFIT2 cells were enriched in highly-infiltrated marrows populated by stressed tumour cells (marrow infiltration: R = 0.55, tumour stress: R = 0.6; Fig. 5a-b, Extended Data Fig. 7h-i). Several other clusters possessed a significant correlation with tumour stress, including positive and negative associations with CD8.Trm and CD4.Tem, respectively (Extended Data Fig. 7i). Teff.IFIT2 cells were not uniquely defined by stress-associated genes (*P* = 0.12, GSEA of stress pathway among Teff.IFIT2 marker genes), arguing against this association representing exposure to similar stressors across T and tumour cells. Additionally, the expression of a smaller set of T-cell effector genes (*CD69*, *TNF*, *IFNG*) followed the same correlations (marrow infiltration: *R* = 0.43, tumour stress: *R* = 0.79, Pearson correlation; Fig.5a, Extended Data Fig. 7j), suggesting a T-cell effector program specifically was associated with infiltration and tumour stress.

Concurrent tumour-associated T-cell memory and effector differentiation suggests a differentiation process between these two states. Clonally expanded Teff.IFIT2 cells enriched in highly-infiltrated marrows populated by stressed tumours expressed markers characteristic of *GZMB*-expressing CD8.TEMRA (Fig. 5c, Extended Data Fig. 7k). This expression was attributable to cells derived from *GZMB*-expressing CD8+ terminal memory clones (Fig. 5c) and clonally expanded Teff.IFIT2 clones predominantly in these *GZMB*+ phenotypes (Fig. 5d, Extended Data Fig. 7k). Therefore, terminally differentiated memory clones accumulating in myeloma are continuous with effector-like T-cells in infiltrated marrows, suggesting these states are linked by a T-cell differentiation pathway.

### Myeloma-associated T-cell differentiation possesses features of anti-tumour immunity

The enrichment of clonal memory T-cells independent of age and viral specificity in myeloma suggests tumour-directed T-cell responses. To investigate this, we identified TCR predicted to bind autologous tumour neoantigens in two patients (see Methods). Expanded TCRs predicted to bind neoantigens mapped to terminal memory and effector-like cells (CD8.TemTerm: odds ratio = 3.2, *P* < 0.001, ISG.IFIT2: odds ratio = 2.5, *P* < 0.001, Fischer’s exact test; Fig. 5h, Extended Data Fig. 8a) and possessed significantly higher expression of the non-viral specificity signature (*P* = 0.027, Wilcoxon test), suggesting tumour antigen-specific may be involved in MM-associated T-cell differentiation.

Tumour antigen-driven T-cell differentiation suggests a degree of tumour immunogenicity. Therefore, we examined the expression of antigen-expression genes in tumour cells. T-cell skewing and non-viral specificity signature expression did not correlate with tumour MHC-associated pathways (Extended Data Fig. 7a, 8b), possibly as these pathways reflect MHC in the context of interferon signalling^25^. We identified *de novo* pathways enriched among each individual patient’s tumour cells (see Methods), revealing MHC and antigen processing and presentation genes were frequently upregulated by individual tumours (*Antigen processing and presentation* pathway, GSEA adjusted *P* < 0.1 in 6 of 16 (37.5%) of tumours tested; Fig. 5f, Extended Data Fig. 8c, Supplemental Table 2). Non-viral specificity signature expression was highest in patient’s whose tumour cells significantly upregulated MHC pathways (*P* = 0.021, Wilcoxon test; Fig. 5g), suggesting the reactivity component of MM-association T-cell differentiation was connected to tumour MHC class I expression and potentially antigen presentation. Finally, in CoMMpass, patients whose tumours highly expressed MHC pathway genes had superior outcome (*P* = 0.031, log-rank test; Fig. 5h), suggesting that T-cell differentiation dynamics associated with high tumour MHC class I expression may influence clinical outcomes.

## Discussion

Recent insights into anti-tumour T-cell immunity have largely derived from studies of solid tumours but a similar understanding in haematological malignancies is lacking. Myeloma has a clinically-defined precursor disease phase lends itself to the study of how T-cell differentiation is altered with disease evolution.

In this study, we curated a large cohort of single-cell data across 11 studies and 234 donors to identify myeloma-specific alterations to T-cells, which we enhanced through the analysis of the TCR repertoire, the BM and PB, and tumour cell transcriptional state. This allowed us to identify the specific features of tumour biology associated with T-cell differentiation in myeloma, independent of natural heterogeneity attributable to tissue localisation and age. We describe two patterns of myeloma-associated T-cell differentiation (Fig. 5i): (1) terminal memory T-cells with features of antigen-specific differentiation and lacking viral-specificity accumulate dependent on serum paraprotein and tumour MHC expression; and (2) effector T-cells are enriched in highly-infiltrated marrows populated by stressed tumour cells. As these two myeloma-associated T-cell subsets are clonally related (Fig. 5c-d), we suggest they represent the differentiation of tumour-reactive clones accumulating alongside anti-tumour immunity through disease evolution (Extended Data Fig. 9).

It will be important to see if other immune subsets possess similar associations. In pursuit of this goal, we make our integrated data available for other researchers.

Our results resolve conflicting reports on the presence of exhausted T-cells in myeloma and a poor history of checkpoint inhibition in this setting^24,40,41^. We show exhausted T-cells are not pervasively enriched in myeloma and are distinct from the more abundant activated CD8+Tem cells which match previous descriptions of PD1-expressing CD8+ T-cells in healthy donors^42,43^. The presence of these “pseudo-exhausted” cells may have led to misidentification of exhausted cells in myeloma patients^40,44^, especially given their enrichment in the bone marrow (Fig. 2g). Additionally, we did not connect an exhaustion phenotype to tumour-specificity (Fig. 4f, 5e), suggesting exhaustion-associated loss of tumour-reactive T-cells may not drive progression as is thought in solid tumours^1^. However, activated CD8+Tem may still be involved in bone marrow pathology, with cells resembling this phenotype possessing negative and positive associations with the response to T-cell engager (TCE) therapy in advanced MM and combination therapy in SMM, respectively^23,45^.

We show myeloma is associated with an enrichment of terminally differentiated clonal memory T-cells. This analysis extends previous reports^14,21,46^ by demonstrating an independence from age and showing concurrent changes to the TCR repertoire and systemic T-cell compartment. These alterations are similar to T-cell immunosenescence changes seen during aging^12^, meaning SMM and MM patients have prematurely aged T-cell compartments (Fig. 3d). Exaggerated T cell aging in precursor conditions may explain the increased risk of infections in these patients^47^, and impede the ability to control tumour growth, facilitating progression. Prior immunosenescence may also predispose individuals to cancer development hence present more frequently in patients^48^. Additionally, this pattern of T-cell differentiation could mechanistically represent anti-tumour T-cell responses. Patient-derived T-cells show evidence of tumour-reactivity in myeloma^49,50^, specifically terminal memory CD8+ phenotypes^51^. This supports our *in silico* evidence that tumour-reactivity may contribute to myeloma-associated T-cell differentiation (Fig. 4g,h, 5e). This suggests that effector T-cells enriched in infiltrated marrows may represent direct tumoricidal T-cell responses. Repeated waves of tumour growth and T-cell control over time may give rise to memory skewing (Extended Data Fig. 9), akin to successive infections giving rise to terminal memory cells through aging^12^. This could explain the lack of exhausted T-cells in myeloma, as T-cell stimulation would be intermittent (dependent on tumour growth) versus chronic, suggesting factors besides T-cell-intrinsic loss of functionality drives progression. However, alternative processes may drive T-cell differentiation in myeloma: inflammation, pervasive in the myeloma marrow^52^, can drive non-canonical memory T-cell differentiation^53^. *In vitro* validation of tumour-specific TCR clones and mapping their phenotype through disease evolution will elucidate the role of T-cell specificity in myeloma.

As T-cell skewing tracked with disease advancement, it may identify donors at risk of early progression. The ability to track this skewing in the PB makes it attractive for immune prognostication (Fig. 3e). However, T-cell skewing was similar in asymptomatic SMM and overt MM (Fig. 3). While this suggests functional anti-tumour responses occur in high-risk precursor conditions, arguing for early intervention with T-cell-dependent immunotherapeutic interventions such as T-cell engager therapy^45^, it may preclude the use of T-cell skewing to identify SMM patients at imminent risk of progression. Additionally, T-cell skewing was more closely associated with serum paraprotein than clinical diagnosis (Extended Data Fig.7g). This may have the potential to enhance existing paraprotein-based prognostication particularly in the rare subset of patients with non-secretory disease^54^. Further work is needed to explain the T-cell skewing-paraprotein association, but we note in addition to paraprotein being an indicator of tumour bulk (thus, total tumour antigen burden or tumour-associated inflammation) that malignant immunoglobulin-derived peptides can serve as immunogenic T-cell epitopes^7,55^. The additional associations we identify between effector T-cells with marrow infiltration and putative tumour-specificity with tumour antigen presentation-associated gene expression may provide additional combinatorial opportunities for integrating immune and clinical metrics into predictive measures of disease risk.

While we were unable to compare T-cells features and tumour genomic classification in our dataset, T-cell skewing was previously shown to be enhanced in hyperdiploid patients^46^. Additionally, further work is needing to directly connect beneficial survival outcomes associated with tumour MHC and stress (possibly death-related) gene expression (Fig. 5h, Extended Data Fig. 2i) with T-cell differentiation and function. Finally, to understand how tumour genomics relates to T-cell differentiation, it will also be important to longitudinally profile neoantigen-reactive responses alongside tumour evolution.

Our results provide a conceptual framework for how T-cells are altered during myeloma disease evolution and highlight the importance of contextualising immune heterogeneity with tumour biology when using immune biomarkers in myeloma.

## Acknowledgements

This work was funded by CRUK, the Medical Research Council, and the UCL/UCLH Biomedical Research Centre. We thank members of the CARDMON, COSMOS and RADAR teams, particularly Sayeh Foroughi, Ambreen Rashid and Grant Vallance, and the UCLH Haematology Cancer Trials Unit. We thank all patients who participated in these studies. Work at the CRUK City of London Centre Single Cell Genomics Facility and Cancer Institute Genomics Translational Technology Platform was supported by the Cancer Research UK (CRUK) City of London Centre Award [C7893/A26233]. We thank the International Myeloma Society (IMS) for funding the presentation of this manuscript in abstract form at the 2022 and 2023 IMS Annual Meetings. We thank the authors of the publications whose data was re-analysed for this study for making their data freely available, particularly David B. Sykes, Samuel S. McCachren, Madhav V. Dhodapkar, Romanos Sklavenitis-Pistofidis, Irene M. Ghobrial, Ola Landgren, Gareth J. Morgan, Saad Z. Usmani, Francesco Maura and Reyka Jayasinghe. We thank the Multiple Myeloma Research Foundation (MMRF), the Perelman Family Foundation.

## Methods

### Primary clinical samples

Bone marrow aspirates from individuals with myeloma or precursor conditions were obtained from patients included in one of four ongoing clinical trials: (1) Defining risk in smouldering myeloma (SMM) for early detection of multiple myeloma (COSMOS), a multicentre, observational UK study in smouldering myeloma (NCT05047107); (2) Risk-Adapted therapy Directed According to Response (RADAR), a randomised phrase II/III trial in newly diagnosed patients with multiple myeloma eligible for transplant (UK-MRA Myeloma XV)^56^; (3) Carfilzomib/Cyclophosphamide/Dexamethasone with Maintenance Carfilzomib in Untreated Transplant-eligible Patients with Symptomatic MM to Evaluate the Benefit of Upfront ASCT (CARDAMON), a phase II trial^57^; (4) Biology of Myeloma, an observational study open to all plasma cell disorder patients treated at University College London Hospitals (Research ethics committee reference: 07/Q0502/17). Bone marrow aspirates from non-cancer controls were collected as a by-product of routine elective orthopaedic surgery (hip or knee replacements) via the UCL/ UCLH Biobank for Studying Health and Disease (Research ethics committee reference: 20/YH/0088). All material was obtained after written informed consent in accordance with the Declaration of Helsinki.

For scRNA-seq experiments, bone marrow aspirates were collected in ethylenediamine-tetraacetic acid (EDTA) and processed within 24 hours of collection. Mononuclear cells (MNCs) were isolated by Ficoll Paque density gradient centrifugation, using SepMate tubes (StemCell Technologies). Freshly isolated BM MNCs were analysed for tumour infiltration by flow cytometry (LSRFortessa, 4 laser 16 color). Cells were stained with the fluorochrome-conjugated antibody CD138 (PE, clone MI15, BioLegend), CD38 (PE-CY7, Clone HB7, biolegend), and a fixable viability dye (eFluor 780, eBioscience). Tumour cell marrow infiltration was determined as the frequency of live BM MNCs cells co-expressing CD38 and CD138 as determined via manual gating (FlowJo v10, BD Biosciences) (Extended Data Fig. 1f).

For CyTOF experiments, MNCs were isolated by Ficoll Paque density gradient centrifugation, using SepMate tubes (StemCell Technologies) and cryopreserved in 90% FBS and 10% DMSO for long-term storage in liquid nitrogen.

### scRNA-seq and scTCRseq sample and library preparation

For newly-generated “T cell−enriched/depleted” samples T-cells were enriched from freshly isolated BM MNCs by magnetic separation using a Pan T-cell Isolation Kit and CD15 MicroBeads (Miltenyi Biotec). After sorting, the T-cell depleted and enriched compartments were pelleted and resuspended in 0.04% BSA in PBS at 10^6^ cells/mL and loaded onto the Chromium Controller (10X Genomics). For newly-generated “CD8-enriched” samples T-cells were enriched using the same protocol with the addition of CD4 MicroBeads (Miltenyi Biotec) and only CD8-enriched samples were loaded. This generated a total of 47 libraries. All samples were processed using the Chromium Next GEM Single Cell 5’ Dual Index Kit (10X Genomics, v2) following manufacturers protocol. T-cell and CD8-enriched samples were additionally processes using the VDJ kit (10x Genomics). The libraries were sequenced by Illumina NovoSeq 6000. Sequencing data was processed with CellRanger GEX and VDJ (v6.0.0) using the GRCh38-2020-A and vdj_GRCh38_alts_ensembl-5.0.0 human reference genomes, respectively. Across samples Cellranger GEX called a median of 6367 cells and Cellranger VDJ a median proportion of 0.76 cells with product V J spanning TRA and TRB pairs.

### Filtering, integration, clustering, and dimensionality reduction of scRNA-seq data

scRNA-seq data were analysed and integrated using the python packages scanpy (1.8.2) and scvi-tools (0.15.2)^58,59^. Gene-barcode matrices for all newly-generated and re-analysed samples were assigned unique sample-specific barcodes, merged, and subset to high-quality cells for integration (minimum unique genes > 200, minimum total counts > 500, total percentage mitochondrial chromosome-encoding transcripts < 10%, total percentage transcripts encoding haemoglobin genes *HBB*, *HBA1* and *HBA2* < 20%). Cells called as doublets by the python package scrublet^60^ (0.2.3) were removed. Samples with < 100 high-quality cells were removed prior to integration.

For integration, we utilised single-cell variation inference (scvi)^59^. A subset of 7000 highly variable genes across batches was calculated on log(x+1) normalised gene expression with the function scanpy.pp.highly_variable_genes(adata, batch_key=”batch”) to identify genes with consistently high inter-cellular variation across different batches. Specific gene groups which can vary between cells for technical (mitochondrial, representing cell stress) or irrelevant biological (immunoglobin and TCR genes, representing lymphocyte clonality) reasons were excluded from highly variable genes to prioritise clustering on phenotype-defining genes. The un-normalised expression of these 7000 variable genes was prepared for scvi using the function scvi.model.SCVI.setup_anndata() with sample batch as the batch key and sample identifier and 10x chemistry as categorical covariate keys. A scvi model was then initialised with the following non-default parameters: scvi.model.SCVI(n_latent=30, n_layers=2, dropout_rate=0.2, gene_likelihood=”nb”). These parameters (number of HVGs, number of latent dimensions and hidden layers, dropout rate) were selected through a parameter sweep focused on minimising batch influence on integrated latent representation and retaining biological identify (data not shown). Minimisation of batch influence was assessed by linear regression of latent dimensions against batch covariates as implemented by scib (https://github.com/theislab/scib). The retention of biological identity was assessed by analysing the separation of CD4+ and CD8+ T-cells (the median log ratio of *CD4*-expressing and *CD8A*-expressing cells closest to zero across clusters). This model was trained for a maximum of 400*(20,000*x) epochs where x was the number of input cells. Integration was first performed on all cells then repeated for just T-cell clusters using 5000 highly variable genes and gene_likelihood=”nb” but otherwise identical parameters.

The latent representation of the trained scvi model was used to create a neighbourhood graph using scanpy.pp.neighbors(adata, n_neighbors=10) for subsequent graph-based clustering using the leiden algorithm. The size of the local neighbourhood (n_neighbors=10) and Leiden clustering resolutions were selected for optimum granularity of biological clusters. Analysis of the latent representation was used as input for creation of a uniform manifold approximation and projection (UMAP, scanpy default parameters) or Minimum-Distortion Embedding using the Python package pymde^61^ (0.1.15). For visualisation of a large number of cells on either UMAP or MDE, the R package scattermore (1.0) was used to created rasterized dot plots.

### Differential expression and pathway analysis of scRNA-seq data

Differential expression between specified conditions was performed using the R package scran (1.26.2) function pairwiseTTests() between specified contrasts with batch as the blocking level for each cell to model for batch effects. Genes were identified as significantly differentially expressed with a false discovery rate (FDR, Benjamini and Hochberg-adjusted *P* value) of < 0.1. Pathway analysis of differentially expressed genes was performed using the R package fgsea (1.24.0) with gene set enrichment analysis of gene sets from BIOCARTA, KEGG and REACTOME databases accessed via the R package msigdbr (7.5.1; Supplemental Table 2).

### Phenotyping gene expression clusters from scRNA-seq data

Cluster markers genes were calculated using log-normalised expression of all genes in a study-aware fashion using the findMarkers function from the R package scran specifying test.type=”wilcox” and batch as the blocking level for each cell. This restricts differential expression comparisons within individual sources and pools the downstream result, meaning no inter-batch comparisons were performed. Marker genes were combined with supervised analysis of the expression of known RNA and protein markers to phenotype clusters. Clusters characterised by expression of known stress-associated genes (for example, *JUN*, *FOS*)^62^ or by co-expression of marker genes for independent phenotypes (for example, T and B cells) were removed. For T-cell cluster phenotyping, clusters lacking expression of *CD3D*, *CD3E* and *CD3G* or both *CD4* and *CD8A* were removed. *De novo* label prediction tools were run with default parameters: Azimuth (https://azimuth.hubmapconsortium.org/) and Celltypst (https://www.celltypist.org/). Manually curated T-cell naïve and cytotoxicity gene signatures were taken from Chu et al.^63^. Gene sets were applied to cells using the R package UCell (2.2.0)^76^.

### Differential abundance analysis of scRNA-seq data

We normalised cell type abundance following a compositional data framework^64^. For each sample, cluster counts were derived and zero values replaced by a Bayesian-multiplicative replacement strategy which preserves the ratios between non-zero clusters, implemented using the R package zCompositions (1.4.0-1) function cmultRepl()^65^, generating zero-imputed pseudo-counts. The centered log-ratio (CLR) transformation was then used to transform pseudo-counts relative to the geometric mean of all clusters in a given sample, implemented using the R package compositions (2.0-6) function clr(). The CLR transformation thus reports cell type abundance relative to the per-sample average seeking to reduce the mutual dependency of proportional data^66^.

For samples from Stephenson et al.^16^, the median age of each age range was used (for example, for the 50-55 group 52.5 was used). Several donors in the dataset were sampled longitudinally, including Oetjen et al.^67^ and Liu et al.^15^. In these cases, only the first longitudinal timepoint was analysed unless otherwise specified.

Normalised cluster abundances were used as input for a combination of intercept-only and additive regression models exploring the relationship between cluster abundance and different conditions (for example, patient group, or patient group and age) as described. Selected comparisons were also performed using a mixed-effect model with an additional random effect term (for sample, study of origin), implemented using the R package lmerTest (3.1-3) function lmer().

### Unsupervised ordination of T cell composition and calculation of exaggerated T-cell aging

Normalised T cell cluster abundance was used as input for PCA using the R function prcomp. Exaggerated T cell aging was calculated by first constructing a linear model examining the relationship between PC1 and age for non-cancer controls only. Next, the age of cancer patients was used to predict PC1 values for each patient in this model. The difference between predicted and real PC1 values (residuals) for each patient was interpreted as the difference between the T cell skewing expected for each patient’s versus their observed T cell skewing, respectively. These residuals-derived values were termed “exaggerated T cell aging”. A patient was considered to have exaggerated T cell aging if their values were greater than zero (or, skewing was greater than expected for their age).

### scTCRseq pre-processing, clonal expansion calculation, T cell subset identification and clustering

TCR paired alpha and beta clones (also termed clonotypes) were defined by CellRanger VDJ (raw_clonotype_id, clonotype_id) by matching shared V and J gene and CDR3 sequences for alpha and beta TCR chains were appended to single cells by matching barcode. For scTCR-seq derived from published data, we used published clone identifiers. Clonal expansion was calculated as the abundance cells labelled with each clone identified in each sample. CD8+ and CD4+ clones were identified by aggregating expression of *CD8A*, *CD8B* and *CD8B2* or *CD4*, respectively, across all cells for each clone. Clone subset was defined as CD4 (N cells CD4 detected ≥ N cells CD8 detected), CD8 (CD8 < CD4), or double negative (DN, CD4 and CD8 both zero; Extended Data Fig. 6a). Repertoire clonality was calculated among each specific subset of cells (such as all T cells or CD8+ memory cells) with a minimum of 100 cells using Simpson’s diversity index^68^. Clusters of TCRs with similar sequence features were identified within a single patient’s alpha or beta chain repertoire using tcrdist3 (0.2.2)^36^ using default parameters. TCR clustering networks were constructed and visualised using the R package igraph (1.4.2).

### Annotation of HLA-matched viral reactivity-annotated TCR clones

HLA genotypes for 19 patients were derived using arcasHLA^69^ ran on Cellranger output bam files (possessorted_genome_bam). All 19 patients were newly-sequenced for this study and therefore a combination of T cell-enriched/depleted and CD8-enriched (Extended Data Fig.1a). arcasHLA was ran on every sample for each patient. HLA genotype for class I and class II HLA was almost entirely identical across samples for an individual. In the rare cases of two different samples possessing different HLA, both predicted genotypes were ignored. Each donor’s repertoire was then compared against the annotated TCR reactivity database VDJdb, IEDB and CEDAR^37–39^ subset to TCRs with annotated reactivity against an epitope from a single human virus: cytomegalovirus (CMV), Epstein Bar virus (EBV), Influenza A, or severe acute respiratory syndrome coronavirus 2 (SARS–CoV–2). TCRs annotated as reactive against more than one human virus were also removed. The viral dataset set was further subset to HLA-matched sequences for each patient’s HLA genotype. A query TCR clone was annotated as putatively viral-reactive if at least one alpha or one beta chain CDR3 sequence perfectly matched a CDR3 annotated against the same virus in the database, and this clone’s paired chain also perfected matched or possessed a highly similar CDR3 sequence to the same virus in the same HLA background. CDR3 similarity was performed as described previously^70^. Briefly, each TCR chain’s CDR3 amino acid sequence was deconstructed into a series of overlapping triplets. Pairwise similarity between two CDR3 was defined as the number of shared triplets normalized to the number of triplets per comparison.

### Identification of predicted neoantigen-reactive TCR clones

Paired whole exome sequencing was performed for two samples as previously described^71^. Nonsynonymous mutations were selected and translated to peptides using a custom script in R. We considered peptides of length 8, 9 or 10 amino acids which contained the altered peptide as potential neoantigens. The immunoglobulin domain as excluded. Intronic mutations or splice variants were not considered. Binding of these to the patient’s HLA class 1 alleles was performed using NetMHCpan (4.1)^72^. Neoantigen binding was deemed significant if the mutant peptide bound the HLA allele with an IC50 of less than 500nm and the wild type had an IC50 of >500nm.

All cases where the peptide-HLA (pHLA) binding criteria were satisfied were considered as potential binding partners for each TCR identified in the sample. TCR–pHLA binding was predicted using TEINet with the authors pretrained models and default settings^73^. The highest scoring pHLA-TCR was considered the most likely binding partner. Among all pHLA–TCR pairs, the phenotype of TCR clones with the highest prediction scores (>0.7) was analysis.

### Identification of malignant plasma cell clones in scRNA-seq

Patient plasma cells were isolated from the clustering of all cells (Fig.1) and patients with < 50 plasma cells were removed. This generated 67,656 plasma cells from 46 patients with a median of 467 plasma cells each (range: 76–13,638). To identify tumour cells among plasma cells, we leveraged the clonal plasma cell origin of myeloma. First, we attached the expression of all available immunoglobin genes to each cell, after removing any gene filtered performed earlier. Then, for each plasma cell, we identified the most highly expressed light variable (kappa or lambda), heavy variable, and heavy constant chains. We next quantified and ranked the abundance of every gene for each chain among an individual’s plasma cells (Extended Data Fig. 2a). Most individuals possessed a single gene for each chain which was expressed by the majority of plasma cells (light variable median 95% range: 31-100%, heavy constant median 96% range: 35-100%, heavy variable median 91% range: 24-100%). The frequency of different light and heavy variable genes among tumour cells matched previously reported frequencies in myeloma^74^, including IGHV3-30 in 3 (6.5%) and IGKV1-39 in 2 (4.3%) patients. We inferred that clonal immunoglobulin expression corresponded to clonal plasma cells and labelled any plasma cell expressing the most highly abundant gene for each chain in that donor as a tumour cell. This method yielded 67,048 predicted tumour cells. Predicted tumour cells uniquely co-expressed clonal immunoglobulin genes (Extended Data Fig. 2b) and expressed genes characteristic of their translocation subgroups (Extended Data Fig. 2c)^75^, suggesting they did represent malignant cells.

### Transcriptional pathway analysis of tumour cells in scRNA-seq data

To analysis tumour cell transcriptomes, we scored tumour cells by a set of pan-cancer transcriptional pathways^25^ using UCell. To compare the expression of individual pathways between patients, we calculated the abundance of cancer cells highly expressing a given pathway as the percentage of cells with expression greater than one standard deviation above the median for each patient’s tumour cells.

### De novo pathway enrichment in malignant cells

To identify novel sets of genes enriched in malignant relative to normal plasma cells, we first isolated each the tumour cells from each patient in turn. Next, we performed differential expression between each patient’s tumour cells only and all other plasma cells not classified as malignant. Differential expression was only performed between cells from the same sequencing batch. This yielded a set of malignant-enriched genes for each patient. Pathway analysis was then performed as described. The pathways significantly enriched among malignant-associated genes in four or more patients were identified (Extended Data Fig.8b).

### Transcriptional pathway survival analysis in CoMMpass

We analysed an association of transcriptional pathways with overall survival in bulk RNA sequencing samples from the CoMMpass cohort of newly-diagnosed MM patients^26^, with RNA sequencing data processed and normalised as described in Bauer et al.^77^. We calculated the expression of each gene pathway by taking of each constituent gene in that pathway, scaling expression between 0 and 1, and taking the average. We assessed the predictive power of each pathway for overall and progression-free survival in patients using the maximally selected rank statistic of the by R package maxstat (0.7-25), and analysed an association between overall and progression-free survival and pathway expression above the maxstat estimated cutpoint using a proportional hazards regression model using the R package survival (3.5-5) with default parameters.

### CyTOF antibody staining, data acquisition and data pre-processing

Details on antibodies are listed in Table S4. Conjugation of the purified antibodies with metal reporters was performed with the MaxPar X8 and MaxPar MCP9 antibody labelling kits (Fluidigm Sciences) according to the manufacturer’s instructions. Frozen bone marrow MNCs or the CD138-negative populations were thawed rapidly at 37°C and resuspended into pre warmed thawing media of RPMI (Sigma-Aldrich) containing 20% FBS, 2mM EDTA (pluriSelect) and 5mg DNase (Sigma-Aldrich). Cell suspensions were washes and filtered to form a single cell suspension. Cells were incubated with 5μM Cell-ID Cisplatin (Fluidigm Sciences) in serum free RPMI for 3 minutes at room temperature (rT) to identify dead cells.

Cells were then washes and incubated with human Fc block (BioLegend) for 10 minutes at rT before being barcoded using a 6-choose-3 Cadmium CD45 Live Barcoding (Fluidigm Sciences). All samples were stained in the same batch. After live cell barcoding, the combined samples were then stained with metal-conjugated antibodies for surface antigens for 30 minutes at rT. After staining, cells were washed with MaxPar Cell Staining Buffer and permeabilised with MaxPar nuclear antigen staining buffer before staining with metal-conjugated antibodies for intracellular antigens. Cells were again washed and fixed using 1.6% paraformaldehyde. Cells were then incubated with Cell-ID intercalator-Ir (Fluidigm Sciences) to stain all cells in MaxPar Fix and Perm Buffer (Fluidigm Sciences) and aliquoted and frozen in cryovials. Stained samples were thawed and washed on the day of acquisition. Cells were acquired on the Helios mass cytometer (Fluidigm Sciences). Data from different days were normalized by using EQ Four Element Calibration Beads (Fluidigm Sciences). Data was debarcoded using the Fluidigm CyTOF software and patient sample fcs files run from different days were concatenated. Before downstream analysis, initial data clean up was carried out using FlowJo. Live CD3+ cells were exported by manual gating on Event_length, Residual, Offset, DNA (^191^Ir and ^193^Ir), live cells (^195^Ir) and CD3 expression (^89^Y).

### Downstream analysis of CyTOF data

CyTOF data were analysed using a custom R pipeline modified from Nowicka et al.^78^. Protein expression data was normalised using the flowCore (2.10.0) logicleTransform() function. Cells were clustered using T-cell markers (all unique proteins shown in Extended Data Fig. 4) using FlowSOM (2.6.0) on a 12 x 12 node self-organising map. This generated 50 putative T-cell clusters. Only clusters expressing CD3 and either CD4 or CD8 were taken retained. One cluster strongly co-expressing all markers was removed as a likely artifact. The remaining clusters were merged to 17 final clusters based on homogeneous co-expression of known T-cell marker genes (for example, CD8+CD45RA+CD45RO-IL7R+TCF7+ were classified as naïve CD8+ T-cells). The expression of T-cell marker proteins for 1000 cells per-sample was used to calculate a UMAP using the R package uwot (0.1.14) with the following parameters umap(expression, n_neighbors=25, metric=”cosine”, spread=2, min_dist=0.1, fast_sgd=TRUE) based on visual separation of clusters.

### Comparison of T-cell clusters across scRNA-seq and CyTOF

First, T cell markers shared between the scRNA-seq and CyTOF datasets were identified. These included (protein/RNA) TBET/*TBX21*, FOXP3/*FOXP3*, “TOX/*TOX*, LAG3/*LAG3*, CTLA4/*CTLA4*, KLRG1/*KLRG1*, PD1/*PDCD1*, EOMES/*EOMES*, CD28/*CD28*, TCF1/*TCF7*, CD69/*CD69*, CD4/*CD4*, CD8/*CD8A*, ICOS/*ICOS*, IL7R/*IL7R*, TIGIT/*TIGIT*, CD25/*IL2RA*, KI67/*MKI67*, GZMB/*GZMB*, HLA-DR/*HLA-DRA*. We also restricted our analysis to scRNA-seq clusters likely to be profiled using our existing CyTOF panel. Therefore, we removed the interferon-expressing ISG.ISG15 and Teff.IFIT2 and invariant MAIT_gdT scRNA-seq clusters. Additionally, we removed the proliferating T cell (Prolif.) scRNA-seq cluster, to avoid a confusion when comparing to the CyTOF CD4.Tm-Prolif. and CD8.Tem-Prolif. clusters, which were also removed. The average expression of each shared marker was scaled within scRNA-seq or CyTOF clusters. The correlation between each cluster’s shared markers was calculated using Pearson correlation (Extended Data Fig.4d-e), with highly correlated clusters inferred to represent the same underlying T cell phenotype.

### Statistical analyses

For comparison of means in box plots, *P* values were calculated by two-sided Wilcoxon test using the R package ggpubr (0.6.0). R and *P* values for correlations were calculated by Pearson correlation. For hierarchal clustering on heatmaps, Euclidean distance was used as the default distance measure.

### Data availability

Published datasets were acquired following the instructions in each original publication. Specifically, data shared through the gene expression omnibus (GEO) can be accessed for Maura et al. under accession GSE161195, Bailur et al. under accession GSE163278, Oetjen et al. under accession GSE120221, Granja et al. under accession GSE139369, Zavidij et al. under accession GSE124310, Kfoury et al. under accession GSE143791, and Zheng et al. under accession GSE156728. Data shared via dbGaP for Sklavenitis-Pistofidis et al. can be accessed under accession phs002476.v1.p1. Data shared online can be accessed for Stephenson et al. (via https://covid19cellatlas.org/), Conde et al. (via https://www.tissueimmunecellatlas.org/), and Liu et al. (via https://explore.data.humancellatlas.org/projects/2ad191cd-bd7a-409b-9bd1-e72b5e4cce81). The integrated single-cell RNA and TCR datasets and cohort information are available online (https://zenodo.org/doi/10.5281/zenodo.11047959). CoMMpass data were downloaded from the MMRF researcher gateway (https://research.themmrf.org). Newly-generated raw sequencing data will be made publicly available and uploaded to the GEO upon peer-reviewed publication. Code to reproduce figures will be made available upon peer-reviewed publication or upon reasonable request.

## Supplemental tables

Supplemental Table 1. Overview of donors included in single-cell RNA sequencing cohort.

Supplemental Table 2. Constituent genes for gene sets and pathways used throughout this study.

Supplemental Table 3. Significantly differential expressed marker genes for T-cell clusters.

Supplemental Table 4. Information regarding CyTOF panel.

Supplemental Table 5. Differential expression results between clones with and without putative viral-specificity annotations.

