## Supplemental Figures for "Tumour-intrinsic features shape T-cell differentiation through myeloma disease evolution"

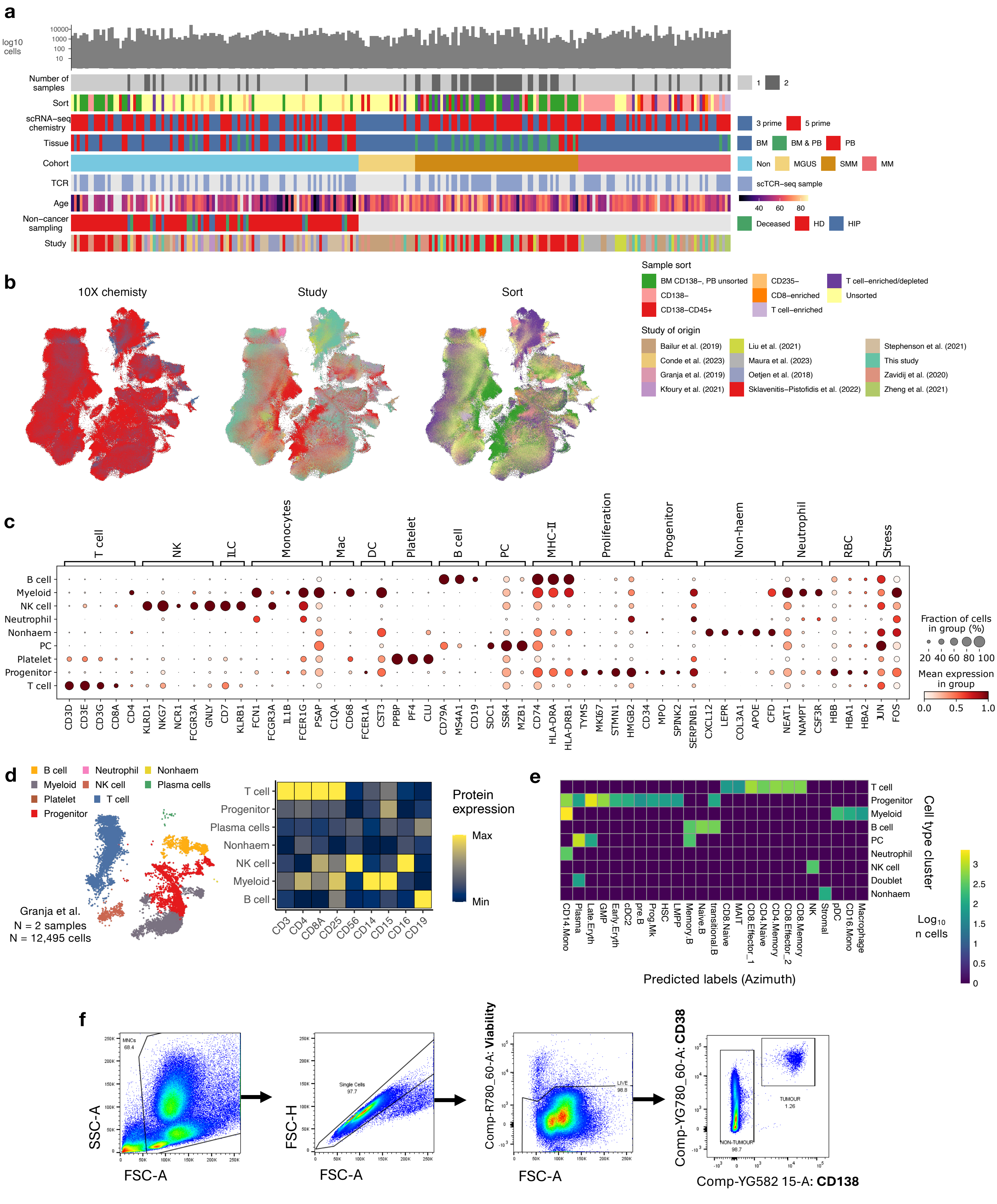


**Extended Data Fig. 1 | Acquisition, integration and immunophenotyping of a single-cell RNA sequencing dataset.** **a**, Heatmap representing an overview of sample metadata for the 237 donors included in the study. BM, bone marrow; PB, peripheral blood; HD, healthy donor; HIP, hip replacement surgery donor. **b**, Visualisation of 1,016,900 single cells in the dataset by uniform manifold approximation and projection (UMAP). The colour of each point represents the indicated covariate matching the legend in (**a**) for that individual cell. **c**, Dot plot of gene expression in all cell type clusters. The mean expression of each gene is scored from low (white) to high (red), with the percentage of cells expressing each gene represented by the size of each point. **d**, Left, UMAP plot showing the cells derived from Granja et al. with individual points (cells) coloured by cell type cluster. Right, Heatmap showing average protein expression derived by cellular indexing of transcriptomes and epitopes sequencing (CITE-seq) in cell type clusters for cells derived from Granja et al. **e**, Heatmap showing the total number of each predicted label (Azimuth, see Methods) for each cell type cluster. **f**, Gating strategy used to calculate tumour cell marrow infiltration in primary bone marrow samples.

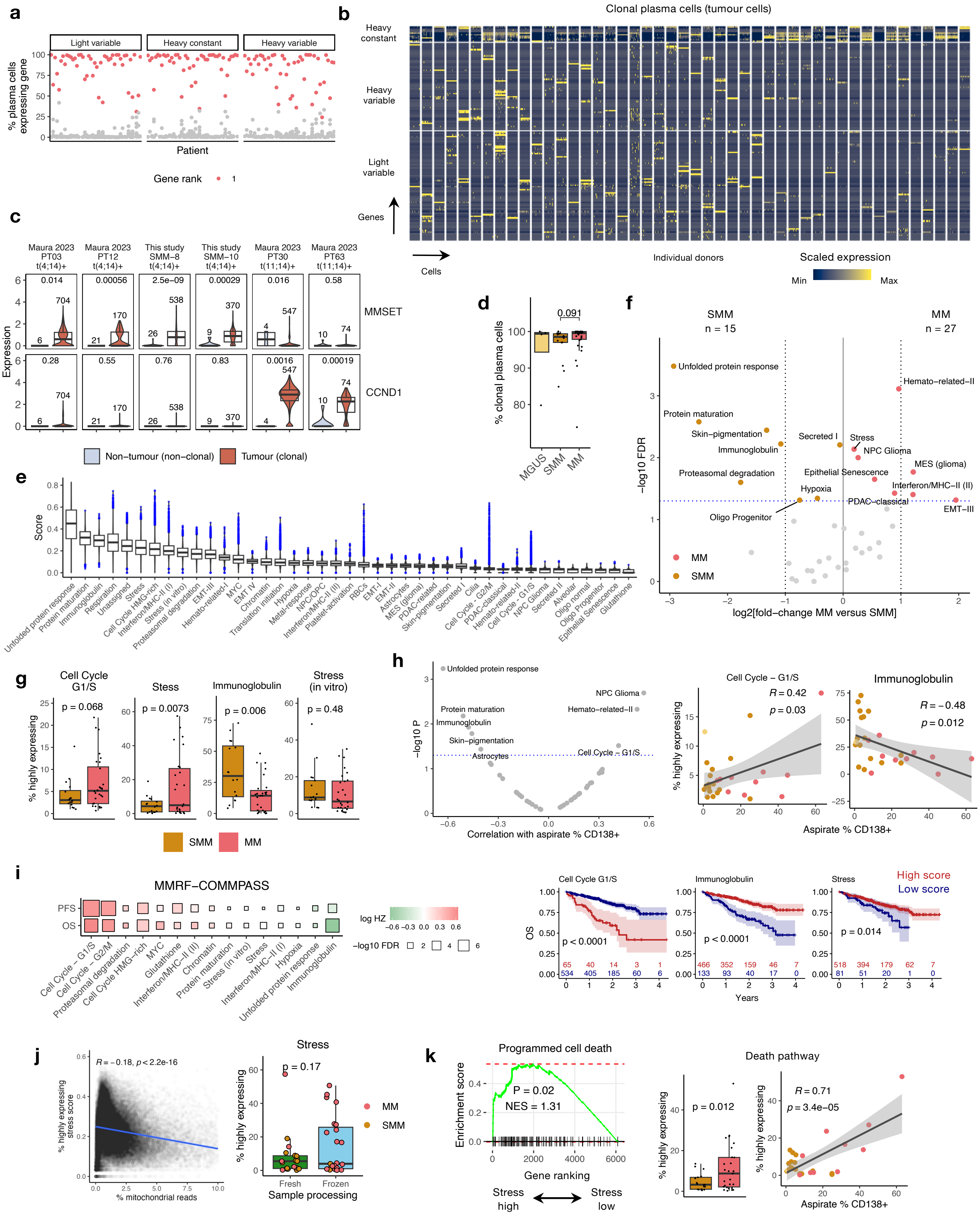

a
Light variable
Heavy constant
Heavy variable
% plasma cells expressing gene
Patient
Gene rank
b
Clonal plasma cells (tumour cells)
Heavy constant
Heavy variable
Light variable
Genes
Cells
Individual donors
Scaled expression
Min
Max
c
Maura 2023 PT03 t(4;14)+
Maura 2023 PT12 t(4;14)+
This study SMM−8 t(4;14)+
This study SMM−10 t(4;14)+
Maura 2023 PT30 t(11;14)+
Maura 2023 PT63 t(11;14)+
Expression
MMSET
CCND1
Non−tumour (non−clonal)
Tumour (clonal)
d
% clonal plasma cells
MGUS
SMM
MM
e
Score
f
SMM n = 15
MM n = 27
−log10 FDR
log2[fold−change MM versus SMM]
g
Cell Cycle G1/S
Stess
Immunoglobulin
Stress (in vitro)
% highly expressing
SMM
MM
h
Correlation with aspirate % CD138+
Aspirate % CD138+
i
MMRF−COMMPASS
PFS
OS
log HZ
−log10 FDR
Cell Cycle G1/S
Immunoglobulin
Stress
High score
Low score
Years
j
% highly expressing stress score
% mitochondrial reads
Stress
Sample processing
Fresh
Frozen
k
Programmed cell death
Enrichment score
Gene ranking
Stress high
Stress low
Death pathway

**Extended Data Fig. 2 | Recurrent tumour cell transcriptional dynamics are associated with progression, local marrow infiltration, and outcome.** **a**, Dot plot showing, for each patient (n = 46), the % of plasma cells expressing each light variable, heavy constant, and heavy variable chain genes. The chain with the highest expression (rank 1) in each patient is shown in red. **b**, Heatmap showing expression of immunoglobulin genes in plasma cells predicted to be tumour cells based on clonal expression of immunoglobulin genes. Each column represents a cell and each row a gene. Columns and rows are split by individual donor and immunoglobulin chain, respectively. A random subset of 50 plasma cells called as tumour cells per donor are shown. **c**, Box and violin plots showing the expression of MMSET (WHSC1 or *NSD2*, upper) and *CCND1* (lower) in plasma cells classified as tumour (clonal immunoglobulin usage, red) or non-tumour (non-clonal immunoglobulin usage, grey) cells in six patients with available genomic translocation information. The number of tumour or non-tumour cells is inset above each box plot. The translocation information (t(4;14) or t(11;14))) is inset below each patient identifier. MMSET is characteristic of t(4;14) positive tumours and *CCND1* of t(11;14) positive tumours. **d**, Box plot showing the proportion of plasma cells with clonal immunoglobulin usage in MGUS (n = 4), SMM (n = 15) and MM (n = 27) patients. **e**, Box plots showing the expression of indicated pan-cancer transcriptional pathways in 67,048 individual tumour cells from all patients. Cells with expression values classified as outliers are shown in blue. **f**, Analysis of the abundance of cancer cells highly expressing each pathway in patients with SMM (n=15) versus MM (n=27). FDR-adjusted *P* values derived by unpaired Wilcoxon test and the $\log_2$[fold-change] for each pathway is represented as a volcano plot. Horizontal dashed line represents the FDR-adjusted *P* value threshold of 0.05. Pathways significantly enriched in either SMM or MM are labelled. **g**, Box plots showing the abundance of cancer cells highly expressing the indicated pathway in patients with SMM and MM. **h**, Left, Dot plot showing *P* values and correlation coefficients between the abundance of tumour cells highly expressing pathways and aspirate % CD138+ values. Horizontal dashed line represents the *P* value threshold of 0.05. Transcriptional pathways with a significant association are labelled. Right, dot plot showing the correlation between the abundance of tumour cells highly expressing the indicated pathways and aspirate % CD138+ values. **i**, Left, Heatmap representing the progression free survival (PFS) and overall survival (OS) associated with the expression of indicated transcriptional pathway in 598 newly-diagnosed untreated multiple myeloma patients enrolled in the ComMMpass trial. Square colour and size represent the log hazard ratio (HZ) and –log10 FDR-adjusted *P* value from univariate Cox regression analysis. Right, Kaplan–Meier curves showing the impact of high (red) and low (blue) scoring of the indicated pathway on OS. *P* values were calculated using log-rank test. **j**, Left, dot plot showing the correlation between the expression of the stress pathway and the percentage mitochondrial read in tumour cells Right, Box plots showing the abundance of cancer cells highly expressing the stress pathway in samples derived from fresh or frozen material. Individual samples are shown as points and coloured by patient group (SMM or MM). **k**, Left, GSEA to show the enrichment of a programmed cell death pathway (M27436) among genes significantly (FDR-adjusted *P* value) enriched in tumour cells highly expressing the stress pathway. Normalised enrichment score (NES) and *P* value from GSEA permutation test is shown. Right, box plot and dot plot showing the abundance of tumour cells highly expressing a death pathway (M14971) in SMM versus MM and in a correlation with aspirate % CD138+. Box plots represent the first and third quartiles around the median with whiskers extending 1.5 times the interquartile range. *P* values shown on box plots were calculated by two-sided Wilcoxon test. R and *P* values for correlations were calculated by Pearson correlation. Correlation shaded regions represent the 95% confidence interval of linear regression slopes.

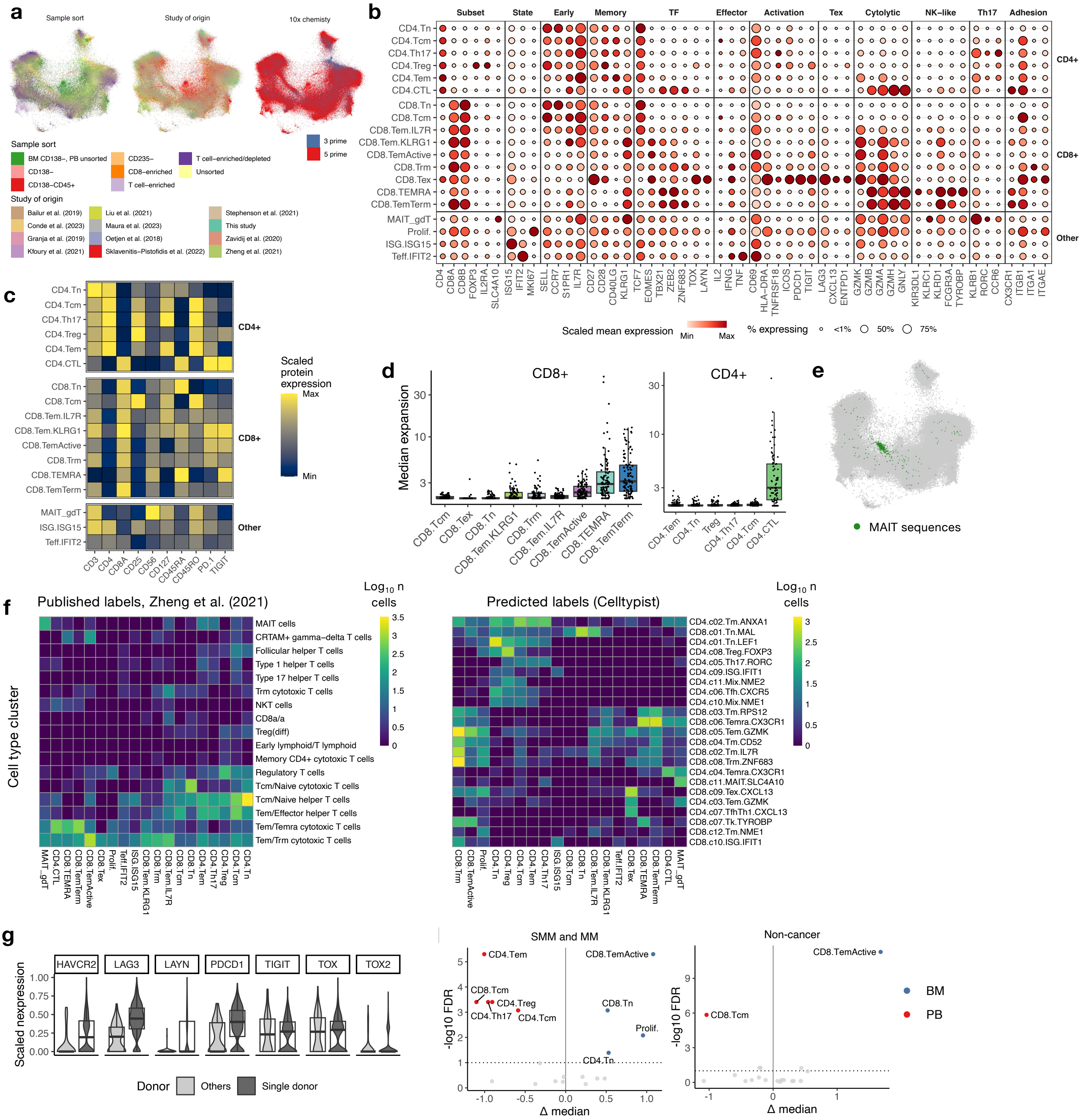


**Extended Data Fig. 3 | The phenotypic and clonotypic landscape of T cells in a large single-cell RNA sequencing dataset of bone marrow and peripheral blood.** **a**, Visualisation of 1,016,900 single cells in the dataset by minimum-distortion embedding (MDE). The colour each point represents the indicated covariate for that individual cell. **b**, Dot plot of gene expression in T cell clusters. The mean expression of each gene is scored from low (white) to high (red), with the percentage of cells expressing each gene represented by the size of each point. Genes are grouped by known associations i.e. T cell memory-associated (columns). TF, transcriptional factor. **c**, Heatmap showing protein expression derived by cellular indexing of transcriptomes and epitopes sequencing (CITE-seq) in T cell clusters for cells derived from Granja et al. **d**, Box plot showing the average T cell receptor (TCR) clonal expansion (calculated as the number of times a unique clone was seen) for all clones in each donors (n = 102). **e**, MDE plot showing the presence of TCR clones with TCR annotated features of Mucosal-associated invariant T cells (MAIT). **f**, Heatmap showing the total number of each published label (Zheng et al.) and predicted label (Celltypist) for each T cell cluster. **g**, Box and violin plots showing the expression of indicated T cell exhaustion associated genes in the CD8.Tex cluster for cells from a single donor (P-20190122, Zheng et al.) and all other donors. **h**, Dot plot showing FDR-adjusted *P* values and difference in the median normalised abundance as a percentage of T cells (Δ median) for T cell clusters and tissue of origin (bone marrow, BM; peripheral blood, PB) for SMM and MM patients (left) and non-cancer controls (right). To account for the fact a subset of samples were taken from donors with both PB and BM samples, *P* values derived by a linear mixed-effects model with donor as a random effect. Box plots represent the first and third quartiles around the median with whiskers extending 1.5 times the interquartile range. *P* values shown on box plots were calculated by two-sided Wilcoxon test.

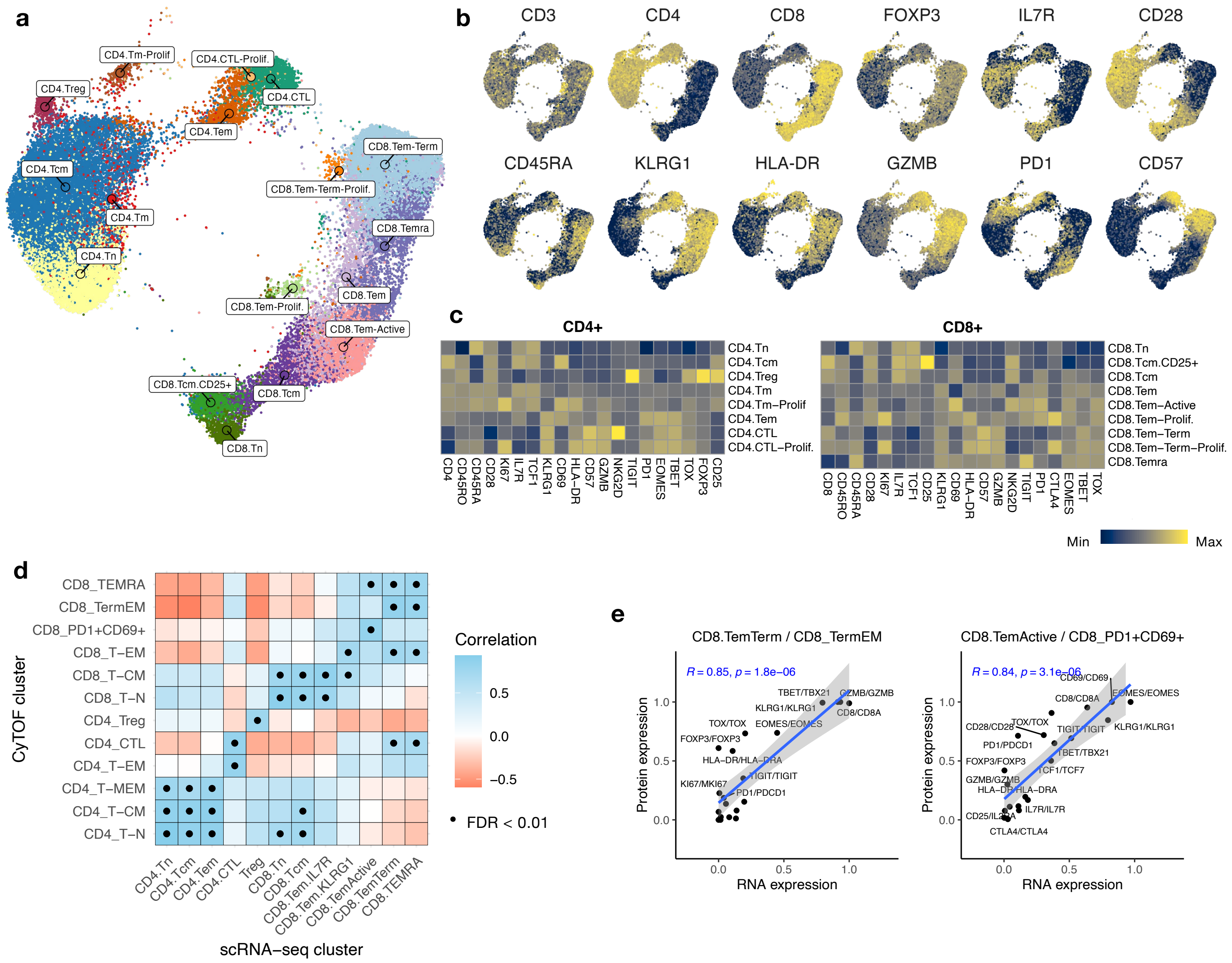

**Extended Data Fig.4 | T cell phenotypes in the bone marrow of SMM and MM patients assed by Cytometry by time of flight (CyTOF). a**, Uniform manifold approximation and projection (UMAP) plot of 100,000 bone marrow T cells derived from SMM (n=9) and MM (n=11) donors. Points (cells) are coloured and labelled by T cell phenotype cluster. **b**, UMAP plots showing the scaled expression of T cell markers **c**, Heatmaps showing the average scaled expression of markers in CD4+ (left) and CD8+ (right) T cell clusters. **d**, Heatmap showing the correlation of scaled average expression of protein and RNA markers in T-cell clusters identified by CyTOF and scRNA-seq (see Methods). Black point represents an adjusted P-value < 0.01. Correlation values and P-values calculated by Pearson correlation. **e**, Dot plots showing the correlation of scaled average expression of protein and RNA markers in indicated CyTOF and scRNA-seq T-cell clusters. R and *P* values for correlations were calculated by Pearson correlation. Correlation shaded regions represent the 95% confidence interval of linear regression slopes.

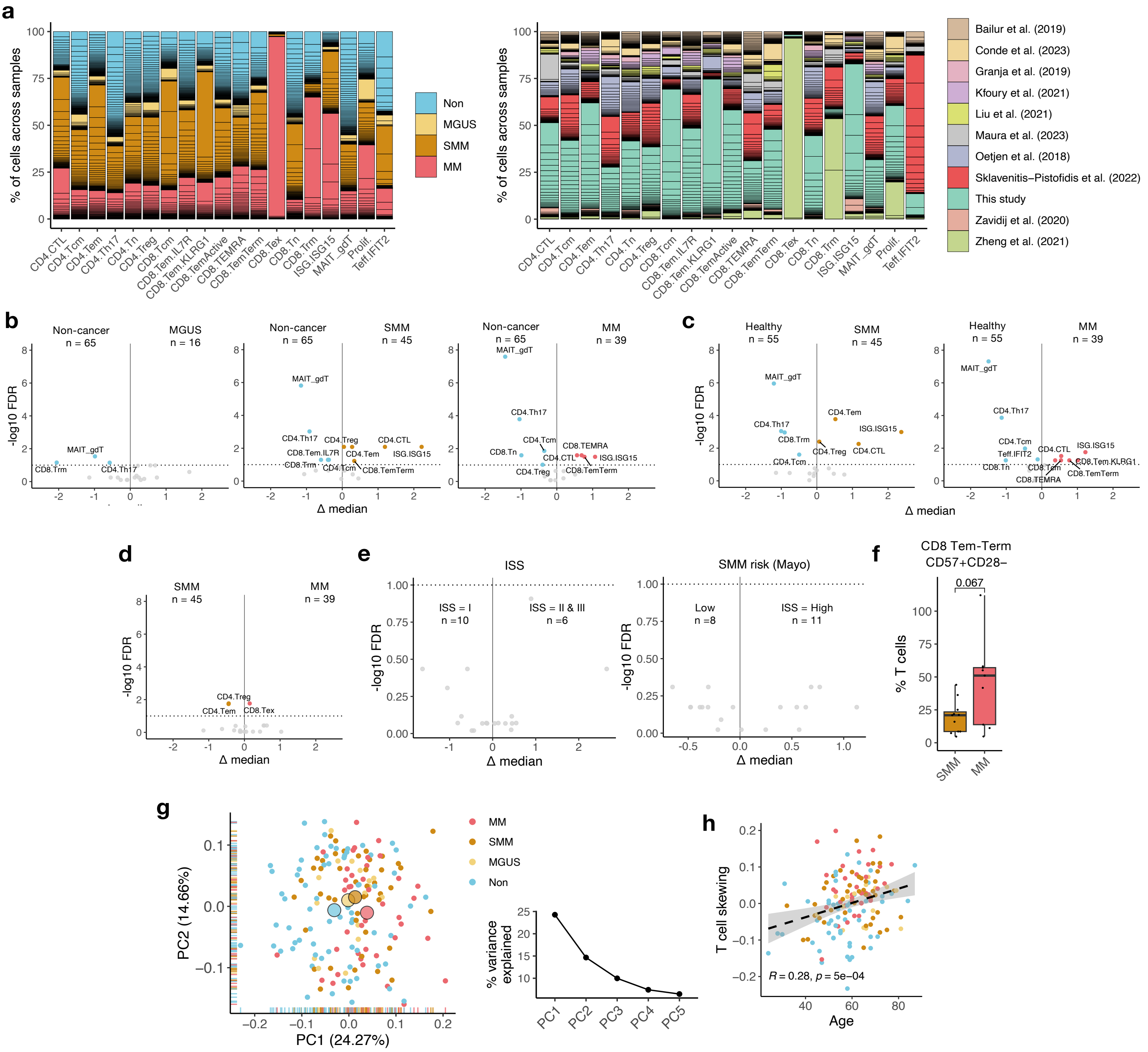

**Extended Data Fig. 5 | Quantification of T cell cluster abundance in non-cancer controls, MGUS, SMM and MM patients. a**, Bar chart showing the percentage of each T cell cluster occupied by individual donors. Individual bar segments represent individual donors and are coloured by cohort (left) or study of origin (right) For example, the CD8.Tex cluster is composed almost entirely of a single MM donor from Zheng et al. **b-e,** Analysis of the abundance of T cell cluster abundance between non-cancer controls and specified patient group (**a**), between true healthy donors (sampled for research) and SMM or MM (**c**), SMM and MM (**d**), and ISS or SMM (Mayo) risk groups (**e**). The number of individuals per group is inset. FDR-adjusted *P* values derived by linear regression with intercept term for age and the difference in the median normalised abundance as a percentage of T cells (Δ median) for each cluster is represented as volcano plots. Horizontal dashed line represents the FDR-adjusted *P* value threshold of 0.1. Clusters significantly enriched in either condition are labelled. **f**, Box plot showing the abundance of CD8.Tem-Term as a percentage of T cells. **g**, Left, Dot plot showing the first two principal components (PCs) calculated using normalised T-cell abundance in all individuals. Large points represent the mean PC1 and PC2 for each group. Right, Scree plot showing the percentage variance explained by the first five principal components calculated on normalised abundance of T cell subsets. **h**, Dot plot showing the correlation between T cell skewing and age (years) in non-cancer controls and MGUS, SMM and MM patients. R and *P* values for correlation calculated by Pearson correlation, with shaded regions represent the 95% confidence interval of linear regression slopes. Box plots represent the first and third quartiles around the median with whiskers extending 1.5 times the interquartile range. *P* values shown on box plots were calculated by two-sided Wilcoxon test.

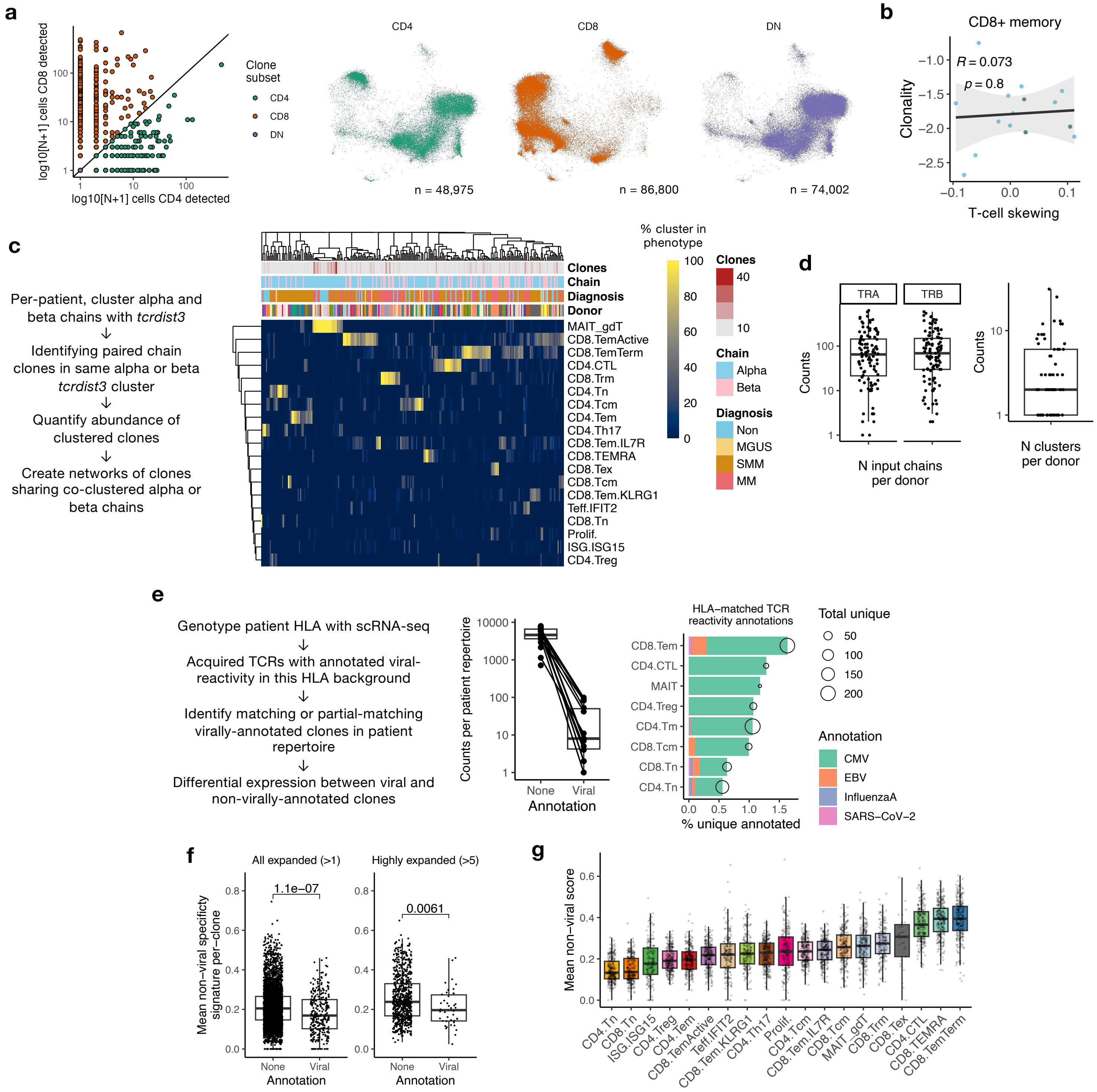


**Extended Data Fig.6 | Analysis of T cell receptor repertoire structure and specificity in myeloma.** **a**, Left, dot plot showing the number of cells with CD8 detected (either *CD8A* or *CD8B1* transcripts non-zero) or CD4 detected (*CD4* non-zero) for all T cell clones (n = 160,258). Clone subset was defined as CD4 (N cells CD4 detected ≥ N cells CD8 detected), CD8 (CD8 < CD4), or double negative (DN, CD4 and CD8 both zero). Right, Visualisation of T cells by minimum-distortion embedding (MDE) coloured and separated by clone subset. The number of clones per subset is inset below. **b**, Dot plots showing the correlation between T cell skewing and clonality ($\log_{10}$ 1/Simpson's diversity) in CD8+ memory clones (CD8+ clones, CD8+ clusters excluding CD8.Tn and CD8.Tcm) from non-cancer controls (n = 15). **c**, Left, Schematic depicting the TCR clustering analysis. Right, Heatmap showing the T cell cluster composition (rows) of 248 individual TCR clusters (columns). The clonal expansion (Clones), TCR chain, clinical diagnosis (Diagnosis), and donor of origin for each individual TCR cluster is shown above the heatmap. **d**, Box plots showing the number of input alpha and beta chains (left) and number of clusters (right) per donor. **e**, Left, Schematic depicting the putative viral specificity clone annotation analysis. Centre, Box plot showing the number of clones viral annotation or no annotation in patient repertoires. Right, Bar chart showing the fraction of annotated TCR specificities against differences viruses across different T cell clusters. **f**, Box plots showing the median expression of the non-viral specificity gene signature among expanded (left) and highly expanded (right) clones with viral or no specificity annotation. **g**, Box plot showing the mean expression of the non-viral specificity gene signature per sample across T cell clusters. Box plots represent the first and third quartiles around the median with whiskers extending 1.5 times the interquartile range. P values shown on box plots were calculated by two-sided Wilcoxon test. R and *P* values for correlations were calculated by Pearson correlation. Shaded regions represent the 95% confidence interval of linear regression slopes.

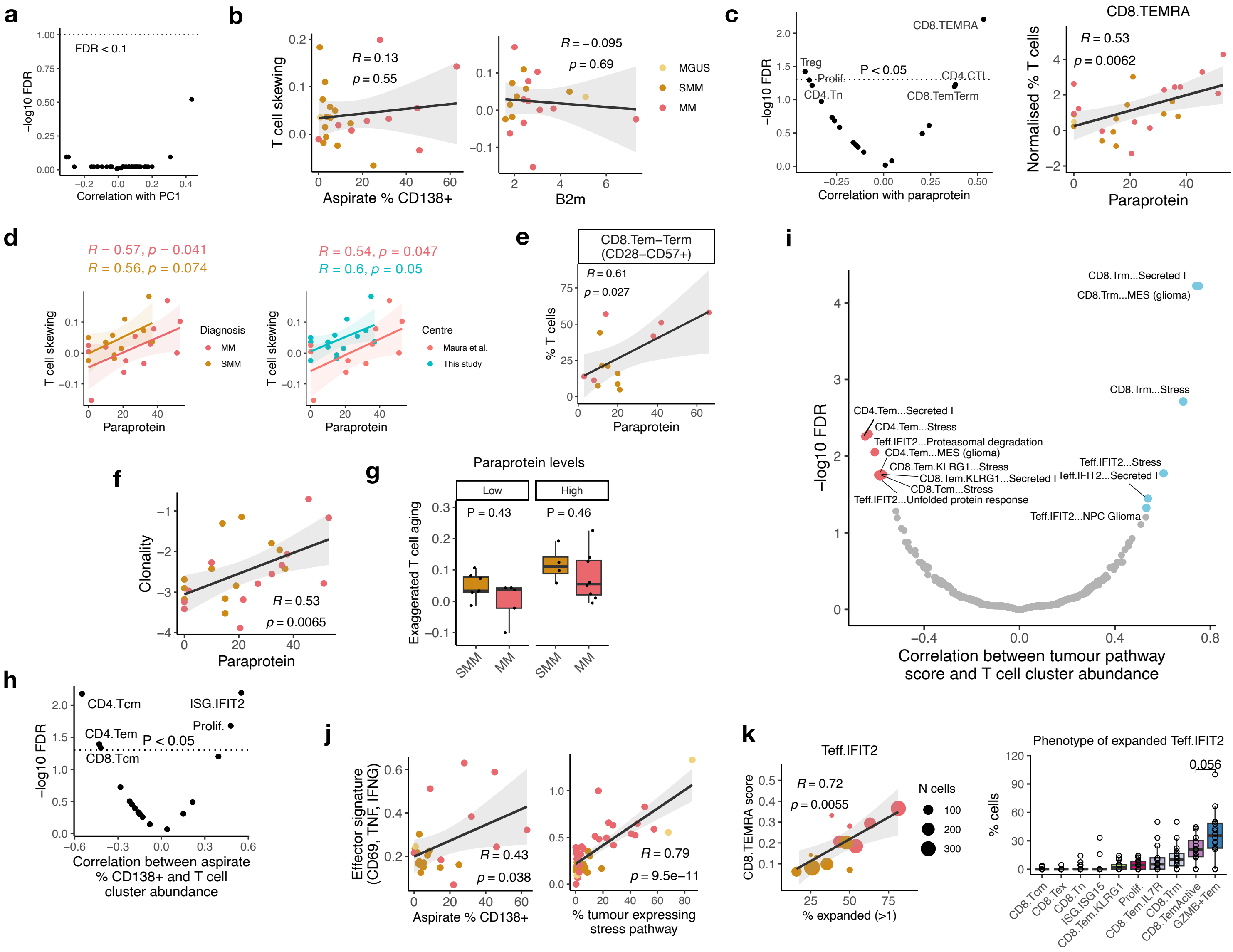


**Extended Data Fig. 7 | Tumour-intrinsic drivers of T cell differentiation in myeloma disease evolution.** **a**, Dot plot showing FDR-adjusted *P* values and correlation coefficients between patient PC1 values and individual pan-cancer transcriptional pathway expression in tumour cells. The dashed line indicates the FDR threshold of 0.1. **b**, Dot plots showing the correlation between PC1 values and the aspirate percentage of CD138+ cells (left) and serum B2m measurements (right). **c**, Left, Dot plot showing FDR-adjusted *P* values and correlation coefficients between patient Paraprotein values and the abundance of T cell clusters. The dashed line indicates the *P* value threshold of 0.05. Clusters which significantly ($P < 0.05$) correlated with Paraprotein are labelled. Right, Dot plots showing the correlation between Paraprotein values and the normalised abundance of CD8.TEMRA. **d**, Dot plots showing the correlation between Paraprotein and PC1 values. Points, curves and correlation values are shown separately for different cohorts (left) and centres (right). **e**, Dot plots showing the correlation between paraprotein values and CD8.Tem-Term cluster abundance for patients from the CyTOF cohort. **f**, Dot plots showing the correlation between TCR repertoire clonality among all T cells and paraprotein values. **g**, Box plot showing exaggerated T-cell skewing (see Methods) in SMM and MM patients with high (> median, 20.7) and low (≤ median) Paraprotein values. **h**, Dot plot showing FDR-adjusted P values and correlation coefficients between T cell cluster abundance and aspirate % CD138+. **i**, Dot plot showing FDR-adjusted *P* values and correlation coefficients between T cell cluster abundance and pan-cancer transcriptional pathway expression in tumour cells. T cell clusters and tumour pathway which were significantly associated are labelled. **j**, Dot plots showing the correlation between the expression of an effector T cell gene signature (*CD69*, *IFNG*, *TNF*) among all T cells with aspirate % CD138+ (left) and the abundance of cancer cells highly expressing the stress pathway (right). **k**, Left, Dot plot showing the correlation between the average expression the twenty most significant CD8.TEMRA marker genes among Teff.IFIT2 cells and the the abundance of expanded (present >1) Teff.IFIT2 clones. Point size reflects the total number of Teff.IFIT2 cells. Right, Box plot showing the percentage each T cell cluster occupied by CD8+ clones which were expanded in the Teff.IFIT2 cluster, excluding Teff.IFIT2. The counts for CD8.TEMRA, CD8.TemTerm and CD4.CTL were combined to GZMB+Tem. Box plots represent the first and third quartiles around the median with whiskers extending 1.5 times the interquartile range. *P* values shown on box plots were calculated by two-sided Wilcoxon test. R and *P* values for correlations were calculated by Pearson correlation. Correlation shaded regions represent the 95% confidence interval of linear regression slopes.

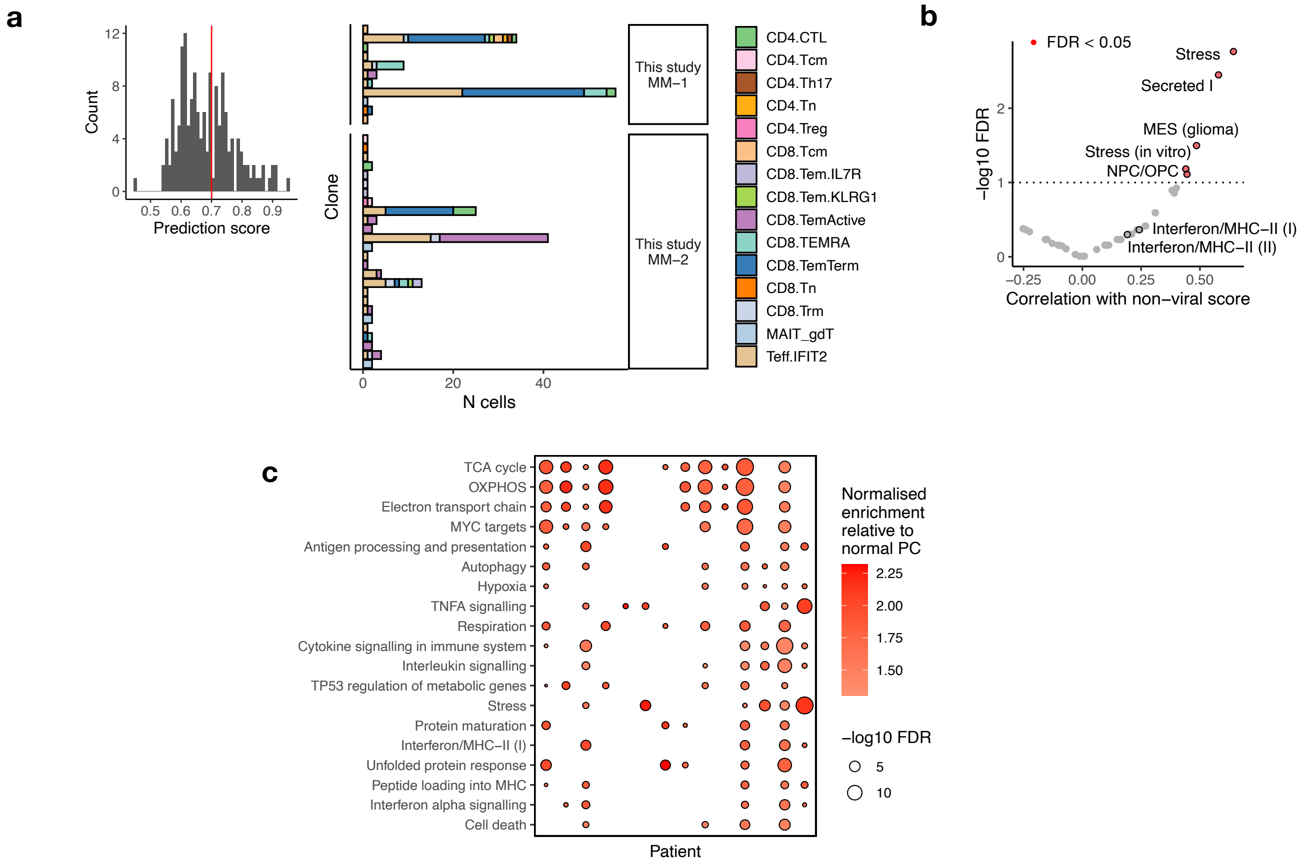


**Extended Data Fig. 8 | Tumour antigen and antigen presentation in myeloma disease evolution. a**, Left, histogram representing the highest neoantigen specificity prediction scores for TCR clones from two patients. The red line represents the threshold for prediction score (0.85) used for selecting clones for further analysis. Right, bar plot showing the cluster composition of clones with predicted specificity score >0.85. The Y axis is split by n = 2 patients. **b**, Dot plot showing FDR-adjusted P values and correlation coefficients between mean expression the non-viral specificity signature in all T cells and pan-cancer transcriptional pathway expression in autologous tumour cells. Pathways with a significant (FDR < 0.05) association and MHC-I and MHC-II pathways are labelled. **c**, Heatmap showing the enrichment of transcriptional pathways on cancer cells relative to non-cancer plasma cells across patients.

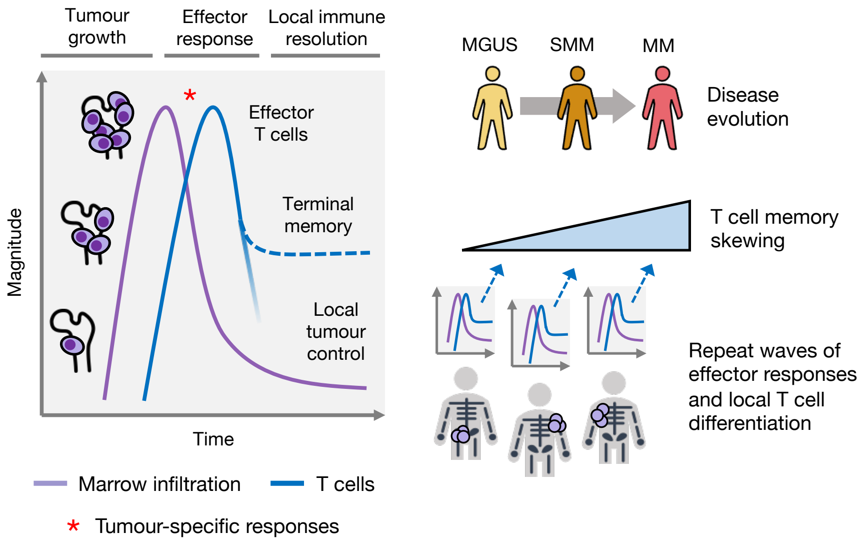


**Extended Data Fig. 9 | Proposal model of T cell differentiation through myeloma disease evolution.** Schematic representing the proposed biological model for T-cell differentiation through myeloma disease evolution. In individual marrow sites, tumour growth leads to increased marrow infiltration. This is mirrored by an expansion of effector T-cells (possibly representing tumour-specific responses), which curtail tumour growth locally in this marrow site. At the resolution of this response, effector T-cells differentiate into memory cells. This process is repeated across marrow sites, over time giving rise to an accumulation of T-cells memory skewing.
